# Decoding Genetics, Ancestry, and Geospatial Context for Precision Health

**DOI:** 10.1101/2023.10.24.23297096

**Authors:** Satoshi Koyama, Ying Wang, Kaavya Paruchuri, Md Mesbah Uddin, So Mi J. Cho, Sarah M. Urbut, Sara Haidermota, Whitney E. Hornsby, Robert C. Green, Mark J. Daly, Benjamin M. Neale, Patrick T. Ellinor, Jordan W. Smoller, Matthew S. Lebo, Elizabeth W. Karlson, Alicia R. Martin, Pradeep Natarajan

## Abstract

Mass General Brigham, an integrated healthcare system based in the Greater Boston area of Massachusetts, annually serves 1.5 million patients. We established the Mass General Brigham Biobank (MGBB), encompassing 142,238 participants, to unravel the intricate relationships among genomic profiles, environmental context, and disease manifestations within clinical practice. In this study, we highlight the impact of ancestral diversity in the MGBB by employing population genetics, geospatial assessment, and association analyses of rare and common genetic variants. The population structures captured by the genetics mirror the sequential immigration to the Greater Boston area throughout American history, highlighting communities tied to shared genetic and environmental factors. Our investigation underscores the potency of unbiased, large-scale analyses in a healthcare-affiliated biobank, elucidating the dynamic interplay across genetics, immigration, structural geospatial factors, and health outcomes in one of the earliest American sites of European colonization.

## Introduction

Determinants of health include a complex interplay of sociodemographic, structural, genetic, and environmental factors that are also contextually dependent on time and geography. Disease risk prediction models and therapeutic paradigms are largely agnostic to many of these important features yet are intended for broad use. Such training datasets often lack the breadth and depth of information and the inherent diversity across features required for equitable applications. The United States populace is highly diverse, marked by complex migration patterns and dynamic social constructs and represents multilevel health contributors. For example, it is widely recognized that the prevalence of diseases is closely linked to individual or neighborhood social deprivation, which further varies across smaller domains and regions^1,2^. Furthermore, these determinants differentially contribute to health outcomes depending on local factors^3,4^.

Contemporary healthcare-associated biobanks represent a new opportunity to discover novel determinants of health and augment translation to clinical care. Such endeavors represent a recent collaborative synergy of large-scale population-based-^5–8^ and local healthcare-biobanks^9–13^. Understanding how DNA sequence variation tracks contemporary and historical population demographics can provide insights on differential disease burdens. For example, rs5742904 (c.10580G>A, p.Arg3527Gln) in *APOB*, a founder pathogenic mutation for familial hypercholesterolemia has significantly higher allele frequency in Old Order Amish people. It substantially explains the increased risk for coronary artery atherosclerosis in this population^14,15^. Important insights related to genetic variations and clinical outcomes may often require profiling diverse populations. For example, the discovery of the association between disruptive variants in *PCSK9*^16,17^ and lower low-density lipoprotein cholesterol in the African population, where these variants are prevalent, facilitated drug development. As another instance, *G6PD* deficiency^18^ was recognized as a prevalent hemolytic disease in Sub-Saharan Africa. Studying diverse populations across a spectrum of diseases is crucial to assess the penetrance of disease-associated monogenic alleles^19^ and polygenic models^20^.

Recent analyses of biobanks in the United States have uncovered the complex genetic structure of Hispanic and/or Latinx groups tracing their origins to the Americas^9,12^. In these efforts, it has been demonstrated that the fine genetic structure within biobanks can identify varying disease risks by capturing both ancestral and social structures, thereby contributing to the advancement of personalized medicine. Separately, recent advances in data size and methodology have enabled us to precisely characterize the complex population dynamics associated with multiple colonization and admixture events^21–23^ However, the interplay across these features, or their interaction with large-scale genetic association studies using whole-genome imputed or sequenced data, remains understudied.

The New England region represents among the earliest European colonization of the United States with sequential ongoing migration from diverse groups. In this study, we examined the genetic variation across New England coupled with sociodemographic, clinical, and environmental/geospatial factors in the Mass General Brigham Biobank (MGBB). By applying a network-based clustering algorithm with newly generated reference dataset, we established fine genetic clusters with subcontinental resolution within MGBB. These clusters exhibited distinct genetic properties, geographic distributions, socioeconomic statuses, and disease risks. In combination with rare and common variant genetic association analyses, we gained further insights into the different disease risks among these populations. Collectively, this study highlights the power of large-scale, unbiased analyses within a healthcare-based biobank to understand the complex interplay between genotypes and phenotypes, paving the way for increasingly personalized interventions.

## Results

### Participant recruitment and Electronic Health Record (EHR) based phenotyping

Since 2010, 142,238 individuals within the Mass General Brigham (MGB) network, the largest healthcare system in Massachusetts, have consented to participate in the MGBB as of May 11, 2023 (Figure 1a, Supplemental Information Figure 1, Supplemental Table 1). Among participants, 99.5% (n = 141,519) consented to re-contact. 56.8% of participants are female (n = 80,851, Figure 1b). Median [interquartile range; IQR] age at consent was 51 [35 – 63] years for female participants and 58 [43 – 68] years for males. Self-reported races were 84.4% White, 4.5% Black, and 3.0% Asian. Self-reported ethnicities were 86.6% non-Hispanic and 2.44% Hispanic (Supplemental Information Figure 2). The participants are primarily cared for at the two flagship MGB hospitals, both located in Boston, MA, and their associated clinics – Massachusetts General Hospital (MGH) and Brigham and Women’s Hospital (BWH, Figure 1c). The biobank data is interlinked with EHRs with phenotype data across the MGB network, as well as notable specialty centers in Boston, MA including the Mass Eye and Ear Institute (MEEI) and Dana-Farber Cancer Institute (DFCI). To generate systemically annotated prevalent/incident outcomes, we extracted International Classification of Diseases codes, Nineth (ICD9) and Tenth (ICD10) revisions, from the EHR and mapped them to PheCodes^24^. We identified 1,577 of 1,860 possible PheCodes with at least one event (Figure 1d). Participants were followed for a median [IQR] of 4.3 [2.5 – 6.0] years after inclusion to MGBB with a median of 12 [5 – 25] incident events per person.

**Figure 1.**
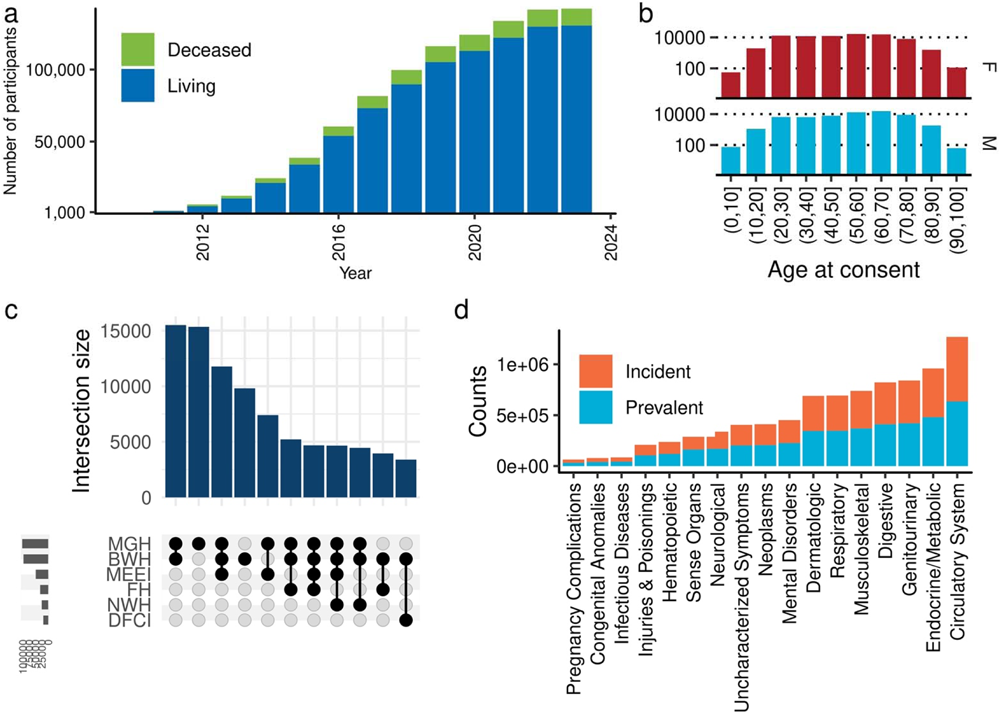
The cohort characteristics of MGBB. **a,** The columns represent the cumulative number of individuals who have consented to the MGBB. Colors indicate the vital status of participants as of May 2023. **b,** Gender/Age at Consent Distribution: The columns represent the distribution of the participants based on gender and age at the time of consent. The individuals older than 100 years at the time of consent are not included in the displayed numbers. **c,** Encounter patterns in the MGB Network. The number of encounters that participants have had within the MGB network. Please note that these encounters include sites where recruitment did not take place. **d,** Distribution of PheCodes based outcomes. The columns indicate the number of outcomes in PheCode-category. Colors distinguish between incident and prevalent cases. MGH, Massachusetts General Hospital; BWH, Brigham’s and Women’s Hospital, MEEI Mass Eye and Ear Institute; FH Faulkner Hospital; NWH, Newton-Wellesley Hospital; DFCI, Dana-Farber Cancer Institute.

**Figure 2.**
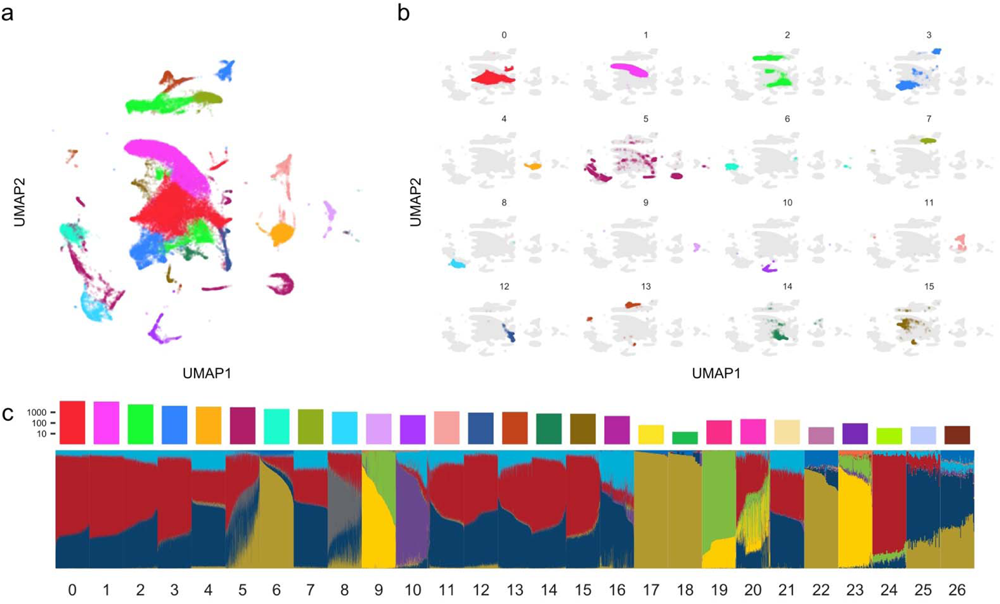
Fine-scale genetic clusters within MGBB. **a,** and **b,** UMAP representation of genetic clusters in MGBB. Each dot represents a participant, with colors indicating distinct genetic clusters identified through graph-based clustering from genetic principal components (Methods). The numbers indicate cluster identification. The color legend and detailed cluster information will be found in Supplemental Table 2 and Figure S2. **c,** Population differentiation in MGBB revealed by ADMIXTURE analysis. The heatmap displays the proportions of ADMIXTURE components (K = 10) within each genetic cluster. The columns at the top of the heatmap represent the number of MGBB participants in each cluster. UMAP, Uniform Manifold Approximation and Projection; MGBB, Mass General Brigham Biobank

### Fine-scale clustering of genetic ancestry in MGBB

Extending beyond traditional low-dimensional projections of continental ancestry from genome-wide data, we utilized high-dimensional principal components (PCs) to achieve greater granularity. Using genome-wide genotyping arrays, we genotyped 53,306 participants in the MGBB. By employing the top 30 genetic PCs and a network-based clustering approach^25^, we identified 30 data-driven distinct ancestral clusters (Figure 2a, 2b, and 2c, Figure S1, Supplemental Table 2). The largest cluster (cluster 0, ordered by sample size) includes 11,875 (22.3%) MGBB individuals. The smallest (cluster 29) includes only one MGBB participant as well as 27/27 reference Sardinian individuals from Human Genome Diversity Project (HGDP)^26^, suggesting the origin of this individual. As such, unsupervised clustering with diverse populations from the 1000 Genomes Project Phase 3 (1KG)^27^ and HGDP reference panels^28^ allowed us to infer the genetic similarity between these clusters and populations worldwide in an unbiased manner.

Cluster 0 was genetically similar to the Western European populations in the reference dataset (CEU [Utah residents with Western or Northern European ancestry] and GBR [British from England or Scotland] in 1KG, French and Orcadian in HGDP, Figure S2). In addition to the cluster 0, we identified eight distinct 1KG+HGDP-EUR-like clusters that cluster with Italian, Russian, Spanish, Adygei, Finnish, Basque, and Sardinian ancestries reflecting known patterns of migration to the Greater Boston Area. We also identified two distinct 1KG+HGDP-AMR-like clusters (cluster 5 enriched with PUR [Puerto Rican in Puerto Rica], cluster 8 with Colombian, Maya, PEL [Peruvian in Lima, Peru], Pima, CLM [Colombian in Medellín, Colombia], MXL [Mexican ancestry in Los Angeles, CA]), four African-like clusters (cluster 6 enriched with African Caribbean in Barbados and African Ancestry in Southwest USA, cluster 17 specific to Nigerian Africans [Esan in Nigeria, Yoruba in Ibadan, Nigeria], cluster 22 specific to Kenyan Africans [Bantu and Luhya in Webuye, Kenya], and cluster 18 with other Western Africans [Mandinka, Mende people in Sierra Leone, Gambian in Western Divisions in the Gambia]), and three East Asian-like clusters (cluster 19 specific to Japanese, cluster 20 specific to Uygur, and cluster 9 with other East Asians). We identified a single large cluster (cluster 10) enriched in South Asian reference populations, however, Hazara and Kalash populations formed distinct clusters.

Even with a diverse reference dataset, eight clusters comprising 9,874 (18.8%) MGBB individuals did not have enrichments of specific populations from the reference dataset. Among these unannotated clusters, seven clusters (4,7,11,13,14,15, and 21) exhibited genetic similarities to 1KG+HGDP-EUR populations. We calculated pairwise Fixation Index (*F*_ST_) values among clusters, and then constructed a phylogenetic tree of the clusters. The observed population differentiation between clusters further corroborates the ancestral relationships but notable distinctions from continental populations and residents in New England (Figure S3a).

We also conducted ADMIXTURE^29^ analyses to infer continuous population structures within these genetic clusters, many of which show similar patterns of structure across increasing numbers of ancestral components (Figure S3b). Using cross validation, ten was the best fit number of components (Supplemental Information Figure 3). We identified two 1KG+HGDP-EUR-like components (distinguished by components 4 and 9). The component 4 was most enriched in the Finnish-like cluster (cluster 24), and relatively enriched in northern European-like clusters (0, 1, and 3) more than the southern European-like clusters (2, 12, and 14). In contrast, the component 9 was enriched in the southern European-like cluster. We also observed a third component included in the European ancestries (component 3). This component 3 is prominent in the Kalash (Indo-European in northwest Pakistan) and other Pakistani reference populations. While this was enriched in southern European-like clusters, it was more enriched in un-annotated European-like genetic clusters 4, 7, 11, 13, and 21 than other annotated European genetic clusters, possibly consistent with Middle Eastern origins as this group is poorly represented in reference datasets.

**Figure 3.**
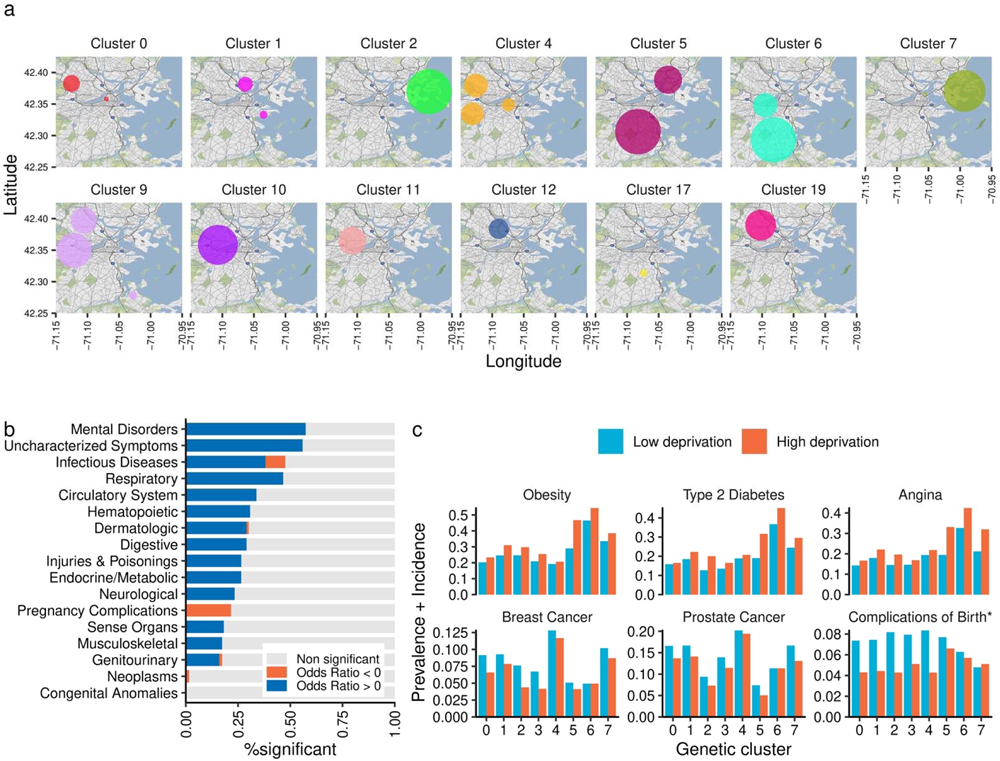
Geospatial distribution, socioeconomic status, and disease risks in MGBB. **a,** Geographical enrichment of genetic ancestries in the greater Boston area. The circle indicates area of significant enrichment of corresponding genetic ancestries. **b,** We tested the association between the socioeconomic deprivation index (SDI) and 1,564 PheCodes based outcomes (Prevalence + Incidence) in 47,070 MGBB participants. The model was adjusted for age, sex, and the first ten genetic principal components. An association was considered statistically significant if *P-*value was less 3.2 × 10^-5^ (0.05/1,564). The color of bars indicated the direction of the effect of SDI (higher SDI suggests higher deprivation). **c,** The disease frequency (prevalence + incidence) by deprivation status and genetic ancestry. The color of bars indicated deprivation status (higher or lower than the median SDI). Complication of Birth, Other and unspecified complications of birth; puerperium affecting management of mother (PheCode 654).

Cluster 4 – the 5^th^ largest cluster in this study (n = 3,514) – is one of such un-annotated European clusters. By comparison of allele frequencies between gnomAD^30^ ancestries and our dataset, we found that cluster 4 has allele frequencies most similar to the Ashkenazi Jewish reference population (Supplemental Table 3). We also observed strong enrichment of skin neoplasms and inflammatory bowel diseases which were previously noted to be enriched in known Ashkenazi cohorts (Figure S4). We also observed significant enrichment of Ashkenazi Jewish founder mutations (e.g., *APC* c.3920T>A, p.Ile1307Lys, *BRCA1* c.68_69del, p.Glu23fs, *BRCA1* c.5266dup, p.Gln1756fs, *BRCA2* c.5946del, p.Ser1982fs)^31,32^ in this genetic cluster (Figure S5). We also observed enrichments of these founder mutations in un-annotated European clusters 11 and 14 suggesting close genetic relationships between these clusters to the Ashkenazi-like cluster 4. In addition to the European-like components, we also identified two different components (components 5 and 7) enriched in the East Asian clusters. One of these components (component 5) was also observed in the Finnish-like cluster which is consistent with previous observations^33–36^.

### Effective population size of ancestry clusters in the Greater Boston area

To characterize the ancestral clusters observed in the MGBB, we estimated the historical transition of effective population size of each cluster using genome-wide genetic data (Figure S6). We conducted IBD-based estimation for effective population sizes (*N*_e_). Our results were consistent with some prior results conducted outside of the U.S. For example, we replicated previously described bottleneck event in Ashkenazi-like population (cluster 4). The lowest *N*_e_ was estimated to be 1,170 (95% CI = 1,100 – 1,270) individuals at 28 generations ago^37^. We observed similar bottleneck events in clusters 11 and 14 around the same generation (minimal *N*_e_ was 4,570 [4,210 – 5,030] in cluster 11 and 32,600 [30,700 – 35,900] in cluster 14) consistent with the aforementioned sharing pattern of Ashkenazi founder mutations with cluster 4. The largest genetic cluster 0 indicates a population bottleneck occurring approximately 12 generations ago. This timeframe coincides with the initial colonization of the Boston area by British settlers. Intuitively, this event is not evident in the British population from the UK Biobank (UKBB) here or in previous studies^38,39^ (Supplemental Information Figure 4), suggesting a unique founder event among British Americans due to colonization. We also observed a significant bottleneck event in the Admixed American populations, specifically in clusters 5 and 8, with a pronounced magnitude in the Puerto Rican-like cluster 5 (minimal *N*_e_ was 11,300 [11,100 – 11,600]). However, we did not observe such bottlenecks for other clusters potentially reflective of more continuous migration.

**Figure 4.**
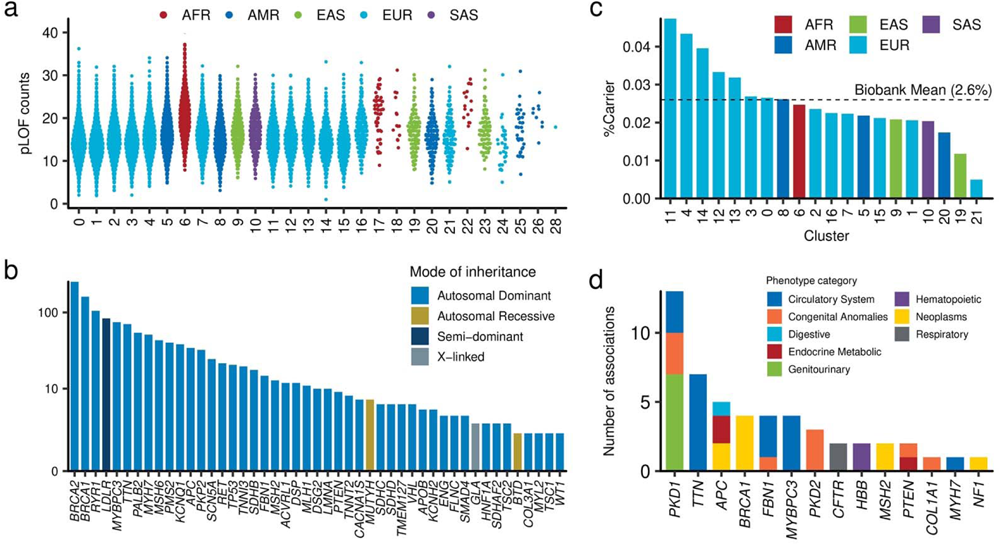
Rare variant identification in the MGBB. **a,** Distribution of the number of protein-truncating variants across genetic ancestries. Each dot represents a participant. The horizontal axis represents the genetic ancestries in the MGBB, while the vertical axis represents the number of protein-truncating alleles in each participant. **b,** Carrier counts of pathogenic/likely pathogenic variants in ACMG actionable genes. The colors of the bars indicate the mode of inheritance of the genes. **c,** Carrier frequency of individuals with pathogenic variants in ACMG actionable genes, categorized by ancestry. The colors of the bars correspond to the continental ancestries. The dotted line represents the average frequency in the MGBB. **d,** Summary of phenome-wide gene burden testing in the MGBB. We conducted exome-wide phenome wide association analysis across 1,454 PheCodes based outcomes in 14,912 genes. The columns indicate the number of significant associations (P < 1.8 × 10^-9^ = 5 × 10^-2^/28,035,307 phenotype-transcript pairs) for designated genes. The color of each column corresponds to the associated PheCode-category. MGBB, Mass General Brigham Biobank; ACMG, American College of Medical Genetics and Genomics; pLOF, predicted loss of function; AFR, African; AMR, Admixed-American; EAS, East Asian; EUR, European; SAS, South Asian.

### Genetic clusters, geographic and socioeconomic factors, and disease risks

We used geospatial scan statistics to understand the geographical structure of MGBB ancestry clusters. We observed 22 statistically significant regions of geographical enrichment among 13 genetic clusters in smaller than 4-km radius areas. We observed distributions of genetically inferred clusters recapitulating colonization histories into the Greater Boston Area (Figure 3a). One example of strong enrichment was observed in the southern area (Roslindale / Mattapan / Dorchester and separately Roxbury) by cluster 6 (ACB [African Caribbean in Barbados] and ASW [African ancestry in Southwest U.S.], expected number 105 and observed number 725, *P* < 1 × 10^-17^). Another strong enrichment is observed northern of Boston (Charlestown / Chelsea) by cluster 5 (PUR [Puerto Rican in Puerto Rico], expected number 180 and observed number 624, *P* < 1 × 10^-17^) in addition to Boston’s South End extending to Roxbury / Hyde Park / Jamaica Plain. We also observed enrichment of Ashkenazi Jewish-like (cluster 4) and East Asian-like clusters (cluster 9) in areas seeded by early founding communities, such as Back Bay / Brookline / Cambridge (cluster 4) and Allston (cluster 9), respectively.

The western European-like clusters cluster 0 and cluster 1 were similar in conventional PC space (Figure S1) and ADMIXTURE analysis (Figure S3b)^29^, but well differentiated by network-based clustering (Figure 2a) as well as geospatially. The CEU/GBR-like cluster 0 was enriched in central areas of the Boston (Beacon Hill) and Cambridge (Harvard Square), representing the earliest sites of British colonization. Cluster 1 (Orcadian-like, tagging northern populations of the British Isles including those hailing from Scotland and Ireland) is enriched in two different geographical locations, including Chelsea and South Boston, where secondary colonization occurred. These different geographical enrichments of cluster 0 versus cluster 1 reflect the distinct histories of these two similar, but distinct genetic European ancestries.

Socioeconomic status was correlated with both genetic ancestry as well as ancestral geographic distributions^1^. Using geocoded location information for each participant in our study, we calculated a Social Deprivation Index (SDI, ranged from 0 to 100, higher SDI indicating greater deprivation) for each participant and associated this with healthcare outcomes (Figure S8, Supplemental Table 4). The distributions of SDI widely differed across genetic ancestries (Figure S7a). Specifically, Cluster 6 (enriched with African Caribbean in Barbados like population) exhibited the highest level of deprivation, with a median [IQR] SDI score of 83 [52 - 94]. Conversely, Cluster 4 (Ashkenazi Jewish-like) had the lowest deprivation, as indicated by a median [IQR] SDI score of 22 [8 - 45].

To systemically identify the associations between socioeconomic status and disease risk in MGBB, we associated SDI with phenome-wide outcomes captured by EHRs, adjusting for genetic ancestries. We found SDI was significantly associated with 400 out of 1,564 phenome-wide outcomes (Bonferroni *P* < 0.05/1,564 = 3.2 × 10^-5^, Figure 3b). Greater SDI was generally associated with increased disease prevalence and incidence (385 out of 400). The strongest SDI-associated PheCodes was with Tobacco use disorder (odds ratio; OR [95%CI]) per one standard deviation (SD) of SDI was 1.54 [1.48 – 1.60], followed by Mood disorders (OR = 1.26 [1.23 – 1.30]), and Depression (OR = 1.26 [1.23 – 1.30]). As represented by these examples, we observed greater risks for Mental disorders, followed by Uncharacterized Symptoms, Respiratory, and Circulatory systems per PheCodes categories, respectively (Figure 3c). However, prostate (OR 0.85 [0.80 – 0.90]) and breast (OR 0.89 [0.84 – 0.95]), cancer had significant/nominal negative associations with SDI (Figure 3b).

Using coronary artery disease (CAD) as an example of a common complex condition, we identified a significant association between SDI and CAD independent from clinical and genetic risk. The association remained significant even after adjustments for clinical risk score (Pooled Cohort Equation, PCE)^40,41^, and polygenic risk score^42^ (PRS, OR_1SD-SDI_ = 1.26 [1.17 – 1.35], OR_1SD-PCE_ = 1.73 [1.65 – 1.81], OR_1SD-PRS_ 1.24 [1.13 – 1.36], in the multivariate model adjusted by the first ten genetic PCs, Figure S7).

### Exome Sequencing in MGBB

Using high-coverage whole exome sequencing in the same group of individuals, we systemically identified rare coding variants in MGBB. There were significant differences in variant distributions across clusters. For instance, the Ashkenazi-like cluster 4 had fewer singleton variants (median [IQR] = 138 [126 – 152] for cluster 4, and 489 [408 – 566] for others). In contrast, there were significantly more singletons in clusters 11 and 14, even though they are closely related to cluster 4 (320 [294 – 361] and 400 [372 – 436], respectively, Supplemental Information Figure 5).

**Figure 5.**
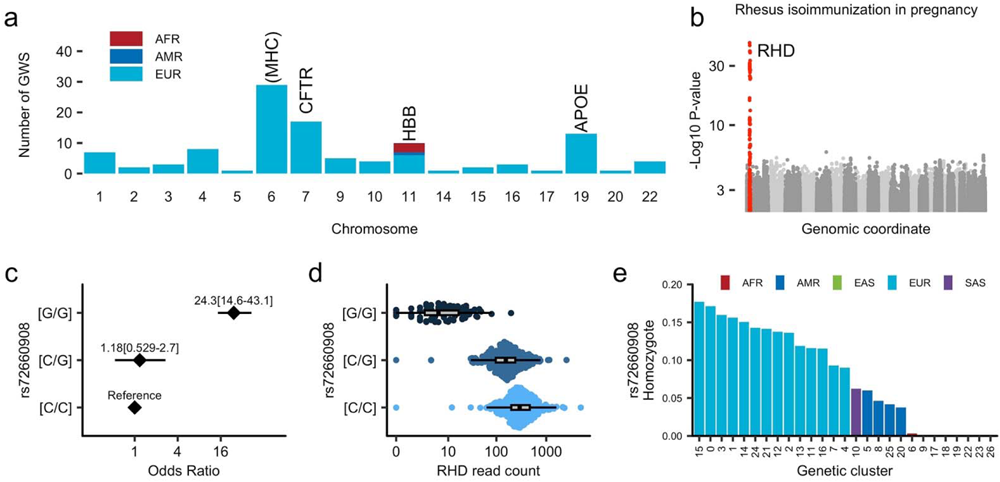
Common variant association study in the MGBB. **a,** Number of associations in the common variant phenome-wide association analysis in MGBB. We conducted associations between common genetic variants (Minor allele counts ≥ 40) and 1,461 PheCodes (Case counts ≥ 60), categorized by continental ancestries (AFR, n = 2,846; AMR, n = 3,756; EUR, n = 44,163). The columns represent the number of significant associations (*P* < 1.6 × 10^-11^) on each chromosome. The color indicates the ancestry in which the association was observed. We annotated chromosomes with more than 10 associations, indicating the representative locus in the chromosome. **b,** Manhattan plot of GWAS for Rhesus isoimmunization during pregnancy in women (n = 23,959). The horizontal axis displays the genomic coordinates from chromosome 1 to chromosome X. The vertical axis represents the strength of association in negative log_10_ *P*-value. The significantly associated variants in the *RHD* locus is highlighted. **c,** Odds ratio for Rhesus isoimmunization during pregnancy by rs72660908 genotypes. The dots and error bars represent the estimated odds ratios and 95% confidence intervals compared to the reference homozygotes ([C/C]). **d,** *RHD* read counts from Whole Blood RNA sequence data obtained from the GTEx dataset. The horizontal axis displays the number of reads aligned to the *RHD* gene, categorized by rs72660908 genotypes. **e,** Frequencies of rs72660908 homozygotes across genetic clusters in MGBB. The horizontal axis corresponds to the genetic ancestries in MGBB, while the vertical axis represents the ancestral frequency of rs72660908 homozygotes ([G/G]). The colors of the columns correspond to continental ancestries. MGBB, Mass General Brigham Biobank; ACMG, American College of Medical Genetics and Genomics; pLOF, predicted loss of function; AFR, African; AMR, Admixed-American; EAS, East Asian; EUR, European; SAS, South Asian; GWAS, Genome-Wide Association Study; GTEx, Genotype-Tissue Expression.

We identified median 15 [12 – 18] rare (Minor allele frequency, MAF < 0.01), high-confidence predicted loss-of-function (pLOF) variants per participant (Figure 4a). The largest number of pLOF variants were observed in African-like clusters (23.5 [20.25 – 26.0] in cluster 22, 21 [19 – 22] in cluster 26, 21 [17 – 24] in cluster 17, 20 [17.0 – 23.0] in cluster 6). The Northern European-like clusters 1 and 0 had the fewest pLOFs (13 [11.0 – 16.0] in cluster 1 and 14 [12 – 17] in cluster 0 and 3). We also identified 1,425 individuals (2.8% of total population) with at least one rare autosomal pLOF homozygous genotype across 761 genes.

We next explored established pathogenic variants (Figure 4b) in MGBB. 2.6% (1,318/50,625) of participants carry a potentially actionable pathogenic/likely pathogenic variant per American College of Medical Genetics and Genomics secondary findings guideline (ACMG SF, version 3.1)^43,44^. These included 6 homozygotes (*TP53*, *LDLR, 4 MUTYH*), and 7 potential compound heterozygotes (2 *BTD*, 3 *MUTYH*, *ATP7B*, and *GAA*). Across genetic clusters, we observed substantial differences in the prevalence of these pathogenic variants (Figure 4c). The highest rate of actionable findings (> 4%) was observed in clusters 11 and 4 despite generally having the lower prevalence of very rare variants. Conversely, non-European clusters generally showed lower rates for annotated actionable variants. Considering higher number of alternate allele-counts in the non-European clusters, the genetic diagnostic rate was significantly lower in non-European populations (Figure S9). Namely, pLOF variants on ACMG SF v3.1 genes found in African and East Asian clusters have significantly lower likelihood of being annotated with a high-quality (more than equal two-stars) pathogenic/likely pathogenic annotation in comparison to the European participants (OR_AFR_ 0.27 [0.18 - 0.41] and OR_AMR_ and 0.48 [0.35 – 0.65], tested by Fisher’s exact test), at least partly related to the underrepresentation of causative variants recurrently observed in non-European groups in the ClinVar^45,46^ database.

To understand the clinical consequences of rare coding variants, we performed exome-wide and phenome-wide association study (PheWAS) across 1,454 PheCodes and 14,912 genes. We did not find substantial evidence of inflation in the test statistics (median Lambda GC_1.0%_ 0.89 [0.86– 0.93] for AFR, and 0.90 [0.87 – 0.93] for AMR, 1.02 [0.99 – 1.05] for EUR, Supplemental Information Figure 6a).

We identified 51 significant associations (*P* < 1.8 × 10^-9^, 0.05/28,035,307 tested phenotype-transcript pairs, Supplemental Table 5) in the burden of rare pLOF and deleterious missense variants with 14 genes, which included 8 ACMG SF v3.1 genes across 45 clinical outcomes (Figure 4d). In addition to the genes associated with known traits, we found significant associations between *PTEN* deleterious variants and increased risk for secondary hypothyroidism. This link was not described by previous rare variant targeted analysis^47,48^ while *PTEN* deleterious variants have been known to be causal for hamartoma syndrome including thyroid cancers and abnormalities^49,50^. Nevertheless, we highlight numerous persistent risk signals from known Mendelian mechanisms of disease in MGBB.

### Genome-wide PheWAS in MGBB

To further explore the relationship between genotype and phenotype in MGBB, we conducted a comprehensive genome-wide PheWAS using ICD code-based PheCodes. We associated over 20 million common variants in African, European, and Admixed populations, which were imputed using the TOPMed imputation reference panel,^51^ with 1,461 PheCodes. Similar to the rare variant burden test, we observed calibrated test statistics overall (median Lambda GC [IQR] were 0.98 [0.95 – 1.00] for AFR, 0.97 [0.93 – 1.00] for AMR, 1.02 [0.99 – 1.03] for EUR, Supplemental Information Figure 6b). We identified 111 associations that reached genome-wide significance (*P* < 5 × 10^-8^/3,048 = 1.6 × 10^-11^, Figure 5a, Supplemental Table 6), including 3 African and 1 AMR associations. We refined the prognosis of identified known low-frequency monogenic variants. For instance, we observed that the variant rs6025 (*F5,* c.1601G>A, p.Arg534Gln; Factor V Leiden) is strongly associated with Congenital deficiency of other clotting factors, including factor VII (OR [95%CI] = 11.56 [9.37 – 14.27], *P* = 3.6 × 10^-68^). Similarly, rs113993960 – a pathogenic variant in *CFTR* (c.1521_1523del, p.Phe508del) – is associated with Cystic fibrosis (OR 14.67 [11.66 – 18.44], *P* = 2.4 × 10^-87^).

Some of these variants exhibited a pronounced recessive effect on the phenotype. A prime example is the variant rs72660908, which is associated with Rhesus isoimmunization in pregnancy (Figure 5b). This medical condition exemplifies recessive inheritance resulting from the deletion of the *RHD* gene. As anticipated, the OR for heterozygotes was not significant (OR_Hetero_ = 1.18 [0.53 – 2.7]) compared to the strong effect observed in homozygotes (OR_Homo_ = 24.3 [14.6 – 43.1], Figure 5c). Recent large-scale sequencing analysis of structural variants^52^ identified high linkage disequilibrium (LD) between rs72660908 and a large deletion affecting *RHD* (R^2^ > 0.99), which we support by strong expression quantitative trait loci (eQTL) effect of rs72660908 on *RHD*^53^ (Figure 5d) and low coverage by exome sequencing in the MGBB (Supplemental Information Figure 7). We also observed a significant enrichment of cases among individuals who were homozygous for rs72660908, with 127 out of 156 cases having the G/G genotype at this locus. As previously reported, individuals with the homozygous alternate allele for rs72660908 have a recessive inheritance pattern in European ancestry populations, but the frequency fluctuates among clusters (17% in cluster 0 and 9% in cluster 4, Figure 5e), and we observed very few copies in AFR/EAS populations.

Another noteworthy example is the association between rs73404549 and sickle cell anemia in the AFR population. This variant is in strong LD with rs334 (*HBB* c.20A>T, p.Glu7Val, HbS), a well-established pathogenic variant for sickle cell anemia. Despite high medical relevance, rs334 was not included in the TOPMed reference panel. We re-evaluated the impact of rs334 using exome sequencing data on sickle cell anemia and clinical red blood cell counts. rs334 showed stronger and more penetrant effect size for sickle cell anemia than imputed rs73404549 (β_rs334_ = 4.14 ± 0.25, *P*_rs334_ = 5.9 × 10^-94^, β_rs73404549_ = 3.05 ± 0.22, *P*_rs73404549_ = 4.4 × 10^-46^) with 14 homozygotes for rs334 and a penetrance rate of 79% (Figure S10a). Additionally, we noted another missense variant rs33930165 in *HBB* (*HBB* c.19G>A, p.Glu7Lys, HbC) – previously shown to confer malarial resistance without sickle cell anemia. We found 5 participants with sickle cell anemia heterozygous for both HbS or HbC with significantly lower red blood cell counts compared to heterozygotes for either genotype (Figure S10b).

## Discussion

In this study, we conducted multidimensional investigations into the structure of a modern healthcare-based biobank based at one of the earliest sites of durable European colonization. We show how expanded immigrant communities in the U.S. often exhibit genetic similarities to contemporary continental populations and reflect common bottlenecks. However, we also observe distinct bottlenecking effects of early colonization and patterns of admixture, and identify populations not well represented in reference datasets. Using geospatial indices, genetic ancestries, and phenome-wide outcome data, we described the architecture of diseases associated with regional socioeconomic factors such as area-level poverty, education level, single parent households, living in rented housing units or overcrowded housing units, living without a care or unemployment^54^. We further leverage rich genotyping and phenotyping to clarify several clinically relevant genetic associations complementing clinical and environmental features. This work advances an overall goal of comprehensively quantifying heterogeneous health determinants that uniquely vary across diverse communities in the U.S.

Leveraging population genetics, we delineated the complex ancestral components present within the Boston area. While our findings align well with prior studies on nationwide cohorts^22,23,38,39^, our research offers further granular insights into the individual-level ancestral histories of the participants, including lifestyle, genetic, and social risk factors associated with the diseases. We used genetic variation to explore the dynamic interplay of migration, expanded colonization sites, and geographic and community variation, aiming to study how social deprivation influences health, independent of the clinical and genetic risk factors. With distinctions from continental level ancestral histories, the complex history of communal-and individual-level factors can be uniquely mapped by healthcare-associated biobanks to uncover novel important drivers of health.

Area-defined SDI improved prediction performance when incorporated into existing clinical^55,56^ and genetic risk stratification models^57,58^ for common complex diseases. In this study, by integrating large scale EHR data and geographical information, we systemically assessed the impact of SDI on Phenome-wide scale across diverse ancestries and drew several clinical implications. First, our systemic assessment suggests that although SDI is a significant contributor to a wide range of diseases, the impacts of SDI are significantly varied across disease domains. For example, while mental and cardiopulmonary diseases are more prevalent among individuals experiencing social deprivation, cancers and congenital diseases are observed almost equally, irrespective of deprivation status. Furthermore, SDI is differentially yet ubiquitously associated with a wide array of health outcomes across various genetic ancestral groups. Finally, although the effect of SDI persisted across various genetic clusters, the varying magnitude of association suggests an interaction between social deprivation and genetic factors as previously suggested^59,60^.

In addition to enabling detailed disease modeling, healthcare biobanks are unique and powerful resources for exploring rare genetic conditions or disease outcomes. First, we identified individuals carrying actionable variants, as defined by a curated database. However, these individuals are predominantly of specific European ancestries, suggesting a bias against non-Europeans in reference datasets, potentially resulting from disparities in clinical genetic testing^19,61^. Using an unbiased genomic scan, our study uncovered several significant associations, which may further refine prognosis within healthcare settings. Furthermore, we confirmed a penetrant association between an upstream variant of the *RHD* gene and Rhesus isoimmunization during pregnancy^13,62^, while also clarifying varied prevalence across diverse communities. Bringing these findings together, we highlight that healthcare biobanks, compared to general population-based biobanks, are enriched with uncommon outcomes, and associated genetic variations, thereby offering an ideal environment to study clinically pertinent scenarios.

Nevertheless, our study warrants several limitations. First, most of our enrollment occurred in tertiary hospitals. While this enabled us to include patients with rare and more severe conditions, the prevalence may not reflect the general population due to inclusion bias as previously described^63^. Second, MGBB participants are centralized in the greater Boston area of Massachusetts, which reflects the geographic location of the two main hospitals of the MGB health system. Communal and geospatial characteristics are likely to vary in other New England regions and more broadly across the U.S. Moreover, while our study provides detailed insights into European populations, the resolution for non-European populations is less robust due to limited sample sizes, reflecting the demography of the included region.

In conclusion, by utilizing a population genetics, we discerned specific ancestral clusters within the MGBB. These clusters reflect the colonization histories of the Greater Boston area and exhibit distinct genetic characteristics and disease susceptibilities. Individual-level clinical and lifestyle risk factors in combination with community context, structural factors, and genetic variation advance disease modeling toward precision medicine initiatives.

## Methods

### Patient recruitment in MGBB and study protocols

MGBB, previously known as Partners Biobank, is an integrated research initiative based in Boston, Massachusetts. It collects biological samples and health data from consenting individuals at Massachusetts General Hospital, Brigham and Women’s Hospital, and local healthcare sites within the MGB network^64^. This repository of samples and data supports researchers aiming to decipher disease mechanisms, enhance personalized medicine, and innovate therapeutic solutions. Since July 1^st^, 2010, the MGBB has enrolled 142,238 participants, and extracted DNA from 88,665 participants’ samples (62.3%). All participants provided written/electronic informed consent for broad biological and genetic research. The study protocol to analyze MGBB data was approved by the Mass General Brigham Institutional Review Board under protocol number 2018P001236. The study protocols to analyze UKBB data was approved under protocol number 2021P002228 and performed under UKBB application number 7089.

### Genotype quality control and imputation

53,306 individuals were genotyped by Illumina Global Screening Array (Illumina, CA) in four batches (13,140 in the 1^st^ batch, 11,649 in the 2^nd^ batch, 5,976 in the 3^rd^ batch, and 22,541 in the 4^th^ batch). Genotypes were called using the Z-call software^65^. After genotype calling, we conducted quality control with the following steps. We re-aligned genotyping probes to the GRCh38 reference genome using the blast software^66^ and extracted probes with perfect-unique match. We removed indels and multiallelic sites and removed variants with high missingness (> 2%) and low minor allele counts (≤ 2). After genotype quality control, we estimated continental level ancestry using the 1KG dataset. We extracted common, high-quality SNPs (missingness < 1%, MAF > 1%) across MGBB and the 1KG dataset. After pruning SNPs, we computed SNP weights for genetic principal component using the 1KG dataset. Then, we projected MGBB participants into the same principal component space using 10 PCs. Using genetic PCs in 1KG dataset as a feature matrix, we trained a K-nearest neighbor model for population assignment (AFR, AMR, EAS, EUR, and SAS) to assign population labels to MGBB participants. With these inferred labels, we calculated Hardy Weinberg disequilibrium for each population and removed variants with *P* < 1 × 10^-6^. Finally, we compared the allele frequency in these populations with gnomAD allele frequency, then removed variants with deviation from ancestry specific gnomAD allele frequency (Chi-square value > 300). These quality control procedures were done by genotyping batch. We took the intersection of the variants in these four batches and generated dataset for imputation. Using the same set of variants, we imputed the genotypes by TOPMed imputation server^67^. We used TOPMed multi-ancestry imputation reference panel (TOPMed r2 panel) including 97,256 reference samples and 308,107,085 variants. Pre-phasing was carried out by Eagle software^68^, and imputation was conducted by Minimac4 software^67,69^. After the imputation, we merged all the four batches by vcftools^70^ and converted to bgen file by PLINK2 software (9 Jan 2023)^71^ for the downstream analysis.

### Exome sequencing and quality control

Exome sequences were performed by on Illumina NovaSeq instruments (Illumina, CA) with a custom exome capture kit (Human Core Exome, Twist Bioscience, CA), with a target of at least 20x coverage at > 85% of target sites. Alignment, processing, and joint calling of variants were performed using the Genome Analysis ToolKit (GATK, version v4.1)^72^ following GATK best practices. The joint called dataset containing all 53,420 individuals processed by Hail framework^73^ for further 1) genotype, 2) variant, and 3) sample quality controls. First, we split the multi-allelic site into biallelic by split_multi_hts function. Following this process, we removed low-quality genotypes and genotypes called by unbalanced allele balance. We consider genotypes that meet the following criteria as missing: For reference homozygotes: total depth (DP) < 10, or Genotype Quality (GQ) < 20. For heterozygotes: DP < 10, Genotype Likelihood for reference homozygote (PL) < 20, (Reference depth + A1 depth)/DP < 0.8, or (A1 depth)/DP < 0.2. For alternate homozygotes: DP < 10, PL < 20, or (A1 depth)/DP < 0.8.

Following genotype quality control, we conducted variant-level quality control. First, we filtered variants in the low complexity region or outside of the target region (broad_custom_exome_v1.Homo_sapiens_assembly38.bed) with 50bp flanks. We excluded i) monomorphic variants and, ii) variants with high missing rate (>10%). Using a quality-controlled variant set, we conducted sample-level quality control. We collected sample QC metric by Hail’s sample_qc function. We implemented five hard filters (percent chimeric reads, percent contamination, call rate, mean depth, and mean genotyping quality, Supplemental Information Figure 8) and four soft filters (number of singletons, Ts/Tv ratio, Het/Hom variant ratio, and Insertion/Deletion ratio, Supplemental Information Figure 9). For soft filters, we obtained residuals of metrics regressing by the first ten genetic PCs and excluded +/-4SD outliers. Finally, using only unrelated quality-controlled samples, we computed Hardy-Weinberg P-values by continental ancestry estimated from genotyping data. Hardy-Weinberg P-values in chromosome X was computed only for Female participants. We excluded variants with ancestry-wise Hardy-Weinberg *P*-values < 1 × 10^-6^ or monomorphic variant. After quality control steps, 7,895,027 variants in 22 autosomes and chromosome X were found in 50,625 individuals remained.

### Relatedness inference

We utilized the pc_relate^74^ function from Hail to adjust for the presence of an admixed population within the MGBB, using 91,615 pruned, common (MAF > 1%) variants that are located outside the major histocompatibility complex (chromosome 6 24,000,000 – 37,000,000 base pair). Among 53,306 individuals, we identified 3,147 pairs with a kinship greater than 0.0884.

### Derivation of genetic principal components

To obtain insights utilizing reference populations, first we combined array genotypes from unrelated MGBB participants with recently generated whole genome sequence datasets from ancestrally diverse populations including 3,381 individuals from 1KG and HGDP^28^. We intersected 495,213 autosomal, non-palindromic variants outside the high LD region with minor allele counts ≥ 10. After merging two datasets, we pruned variants by PLINK2 software^71^ with –indep-pairwise option 1000 100 0.2 resulting in 257,754 variants. Using these genotypes, we derived the weight for each variant for PCs excluding related samples. Using derived weights, we calculated 30 PCs for all the individuals from MGBB, 1KG, and HGDP which were used in subsequent analysis.

### Fixation index

Pairwise Fixation indices (*F*_ST_) were computed among in MGBB-, 1KG-and HGDP-populations using PLINK2 software. The phylogenetic tree was constructed neighbor-joining method^75^ implemented by ape R package^76^.

### ADMIXTURE analysis

Using PCs derived above, we conducted admixed component analysis using ADMIXTURE software (version 1.3.0)^29^. We optimized the number of admix component K from 1 to 20 and found that K = 10 showed the least cross-validation error (Supplemental Information Figure 3).

### Genetic ancestral clustering

To derive fine-scale genetic clusters in the population, we conducted Graph-based clustering which is frequently used in single-cell RNA-seq clustering analysis implemented in Seurat software (version 4.1.0)^25^. Though Seurat is primarily tailored for single-cell RNA seq data analyses, we leveraged its robust clustering capabilities for genetic ancestry clustering. Using the first 30 PCs derived above, we constructed a nearest-neighbor graph and classified individuals into distinct clusters using the Louvain algorithm, a default clustering approach in Seurat with resolution parameter of 0.2. As Seurat identified the clusters in an unsupervised mode, we used individuals from the 1KG or HGDP as a “spike in” positive controls (true labels).

### Effective population size estimation

To estimate the effective population size using haplotype sharing information, we used IBDNe in combination with the hap-ibd. First, we phased the genotypes of unrelated MGBB participants with SHAPEIT software (version 4.2). Then, using hap-ibd software (version 1.0, 15Jun23.92f)^77^, we calculated IBD sharing, and this output was fed into IBDNe software (version 23Apr20)^78^ to determine the effective population size for each ancestry. To compare the effective population size trajectories in British population in UK and MGBB, we computed effective population size in down-sampled, unrelated UKBB European-like population to the same sample size as MGBB British-like population (n=11,508), using microarray-based genotypes.

### Variant annotation

We annotated WES data using the VEP software (version 107)^79^, supplemented with the Loftee^30^ and dbNFSP^80^ plugins. The “—pick” option was enabled to prioritize the canonical transcript. Additionally, in-silico predictions from dbNFSP (version 4.2) were employed to prioritize missense variants.

### Pathogenic variant annotation

We downloaded ClinVar database^45,46^ on Aug 16, 2022, and annotated all the variants identified by exome sequence using snpEff software (version 5.0e) ^81^. We identified 536,729 variants registered in the ClinVar Database overall. To identify the carriers of pathogenic/likely pathogenic variants in the ACMG SF v3.1 actionable genes^43,44^, we only used variants with review status “reviewed_by_expert_panel”, “criteria_provided,_multiple_submitters,_no_conflicts”

### Polygenic risk score

Using the PRS-CS software^82^, we determined posterior weights for 1.2 million hapmap3 SNPs from a previous CAD GWAS^42^, which did not include the MGBB/UKBB population. Given our study’s predominant European population, we utilized the European reference panel provided by the PRS-CS authors. Our model training and derivation of posterior weights incorporated parameters phi ranging from 1 × 10^-1^, 1 × 10^-2^, 1 × 10^-3^, 1 × 10^-4^, and 1 × 10^-5^. With these weights, we determined the CAD-PRS for both UKBB and MGBB populations. From the UKBB results, we identified the optimal parameter phi as 1 × 1 × 10^-3^ and applied this PRS in the MGBB analysis.

### Disease Phenotyping

We obtained patient data from the electronic health record system within the MGB network. We specifically extracted ICD9 and ICD10 codes assigned to each patient. To enhance the interpretability and powered analysis of the disease outcome, we employed the PheWAS R package (version 1.2)^83^ to map these codes to corresponding PheCodes^24^. The PheWAS package utilizes a comprehensive catalog of PheCodes (https://phewascatalog.org/phecodes). This mapping process facilitated a more standardized and consistent representation of the patient’s conditions for subsequent analyses. To determine the prevalence or incidence of diseases, we considered the date of blood draw for genotyping as the reference date. By aligning with the corresponding date of PheCodes occurrences, we identified the prevalent or incident outcomes related to the date of enrollment in MGBB.

### Clinical risk, genetic risk, and social risk for CAD

For the CAD analysis, we calculated the 10-year Atherosclerotic Cardiovascular Disease (ASCVD) risk scores based on the PCE using the *PooledCohort* R package^39,40,84^. The PCE accounts for sex, race, age, total and HDL cholesterol, systolic blood pressure, antihypertensives prescription, current smoking, and prevalence of diabetes mellitus. For missing values, we performed multiple imputation by chained equations using the *mice* R package^85^, using enrollment age, sex, and race as predictors. Among participants without prior CAD, the first post-enrollment CAD incidence was ascertained based on relevant ICD-9 and ICD10 codes from in-hospital records. We assessed the individual association of 10-year ASCVD risk, CAD-PRS, and SDI with incident CAD based on logistic regression.

### Exome-wide burden test

We conducted a rare variant aggregation burden test implemented in Regenie software (version 3.2.5)^86^. We generated masks comprised of predicted loss of function (high confidence by Loftee software^30^) and damaging missense variants predicted by > 90% of 29 in-silico prediction programs included in dbNFSP (version 4.2)^80^ with MAF < 0.01.

### Genome-wide, phenome-wide association study

We conducted single variant PheWAS using imputed genotypes and 1,461 PheCodes. We applied mixed model approach implemented in Regenie software (version 3.2.5)^86^. Null model was fit using pruned common variants derived from microarray-derived genotypes (MAF > 1%, pruned by PLINK2 software^71^ with option –indep-pairwise 1000 100 0.9). The analyses were conducted ancestry wise. For sex specific endpoint, only male or female were included in the analysis. The genome-wide significant threshold was set at *P* = 1.6 × 10^-11^ dividing conventional genome-wide significant threshold 5 × 10^-8^ by 3,048 tested phenotypes across three ancestries. To define the associated loci, we added the flanking region (± 500,000 base-pairs) for all the variants with genome-wide significance (P < 1.6 × 10^-11^) and merged all overlapping regions.

### PheWAS Inflation statistics

We estimated Lambda GC (observed chi-squared value divided by expected value) i) at top 1.0 percentile of the test statistics for rare variant burden PheWAS, and ii) using HapMap3 SNPs for common variant PheWAS. For common variant PheWAS, LD score regression^87^ was additionally performed to estimate intercept using ldscr R package (https://github.com/mglev1n/ldscr).

### Geocoding

The participants’ current address data was geocoded using the DeGAUSS framework^88^, a collection of geospatial tools designed for cleaning and formatting geographic data. This process converts the address information into standardized spatial data, specifically latitude and longitude coordinates. Our analysis focused on participants residing in Massachusetts. We excluded 1) Participants whose addresses were located outside of the state of Massachusetts, 2) Participants for whom the geocoding process failed. We successfully geocoded 48,369 individuals with genotype data.

### Area-based deprivation score index

For individuals whose addresses were successfully geocoded, we proceeded with the following steps: We assigned each individual’s address to a corresponding U.S. Census tract. Census tracts are small, relatively stable geographic areas that are defined by the United States Census Bureau. They are designed to be relatively homogeneous units with respect to population characteristics, economic status, and living conditions. B) We then merged this Census tract-level data with SDI (2018 SDI, downloaded from https://www.graham-center.org/maps-data-tools/social-deprivation-index.html)^54^. SDI is a composite measure of area level deprivation based on seven demographic characteristics collected in the American Community Survey (ACS) and used to quantify the socio-economic variation in health outcomes. The final SDI is a composite measure of seven demographic characteristics collected in the ACS: percent living in poverty, percent with less than 12 years of education, percent single-parent households, the percentage living in rented housing units, the percentage living in the overcrowded housing unit, percent of households without a car, and percentage non-employed adults under 65 years of age. This approach allows for a detailed, regional census tract-level analysis of the social conditions experienced by the study participants.

### Spatial enrichment analysis

We utilized the Bernoulli model in SaTScan^89^. Under this model, individuals belonging to a specific genetic cluster were treated as “cases,” while all other individuals were treated as “controls.” This model compares the rates of cases in different areas to determine if the rate of cases inside the potential cluster area is significantly different from outside. To avoid detecting overly large and potentially less meaningful clusters, we limited our scan by setting the maximal diameter of the spatial cluster window. Specifically, we restricted this to a maximum radius of 4 kilometers.

## Data Availability

Genotyping and exome sequencing data for 13,500 participants from the MGBB are available in dbGAP (https://www.ncbi.nlm.nih.gov/projects/gap/cgi-bin/study.cgi?study_id=phs002018.v1.p1). Additional MGBB data were accessed under institutional review board protocol for this current study and are not publicly available due to restrictions on the data. The summary statistics for phenome-wide common/rare variant association analysis and the allele frequencies of genetic clusters will be publically available upon the publication.

## Acknowledgements

### Funding sources

S.K. is supported by Japan Society for the Promotion of Science (202160643), Uehara Memorial Foundation, and National Institute of Health (NIH), National Heart Lung and Blood Institute (NHLBI, K99HL169733). K.P is supported by the MGH Executive Committee for Research Fund for Medical Discovery. S.J.C. is supported by a grant of the Korea Health Technology R&D Project through the Korea Health Industry Development Institute (KHIDI), funded by the Ministry of Health and Welfare, Republic of Korea (HI19C1330). S.M.U. is supported by NIH National Human Genetics Research Institute (NHGRI, T32HG010464). R.C.G. is supported by NIH (HG009922, OD026553, HL143295, TR003201). B.M.N. is supported by R37MH107649. P.T.E. is supported by grants from the NIH (1R01HL092577, 1R01HL157635, 5R01HL139731), from the American Heart Association (18SFRN34230127, 961045), and from the European Union (MAESTRIA 965286). J.W.S. is supported by OT2OD026553, U01HG008685, MH118233. M.L. is supported by grants from the NIH (OT2OD002750, R01HL143295, U01HG008685, U01TR003201, and U24HG006834). E.K. is supported by grants from the NIH (OT2OD026553, U01HG008685, P30 AR070253, OT2HL161841). A.R.M. and Y.W. are supported by NIH, NHGRI (K99/R00MH117229 to A.R.M.) and by European Union’s Horizon 2020 research and innovation program under grant agreement 101016775. A.R.M. is also supported by U01HG011719. P.N. is supported by grants from the NHLBI (R01HL127564), and NHGRI (U01HG011719).

### Author contributions

R.C.G., M.J.D., B.M.N., P.T.E., E.W.K., A.R.M., and P.N. conceptualized this project. S.K., K.P., S.J.C., and E.W.K. curated phenotype data. S.K., and M.S.L. curated genotype data. S.K., M.U., and S.M.U. analyzed data. S.K., Y.W., P.T.E., E.W.K., A.R.M., and P.N. interpreted data. S.K., A.R.M., and P.N. prepared the initial draft. Y.W., K.P., M.U., S.J.C., W.E.H., R.C.G., M.J.D., B.M.N., P.T.E., J.W.S., E.W.K., A.R.M., and P.N. provided critical review and edits for the manuscript. S.H., W.E.H., R.C.G., M.J.D., B.M.N., P.T.E., E.W.K., A.R.M., and P.N. supervised the project. S.H., W.E.H., P.T.E., J.W.S., M.S.L., and E.W.K. managed the project administration. P.T.E., J.W.S., M.S.L., E.W.K., and P.N. obtained funding for the project.

## Supplemental Figure Legends

**Figure S1.**
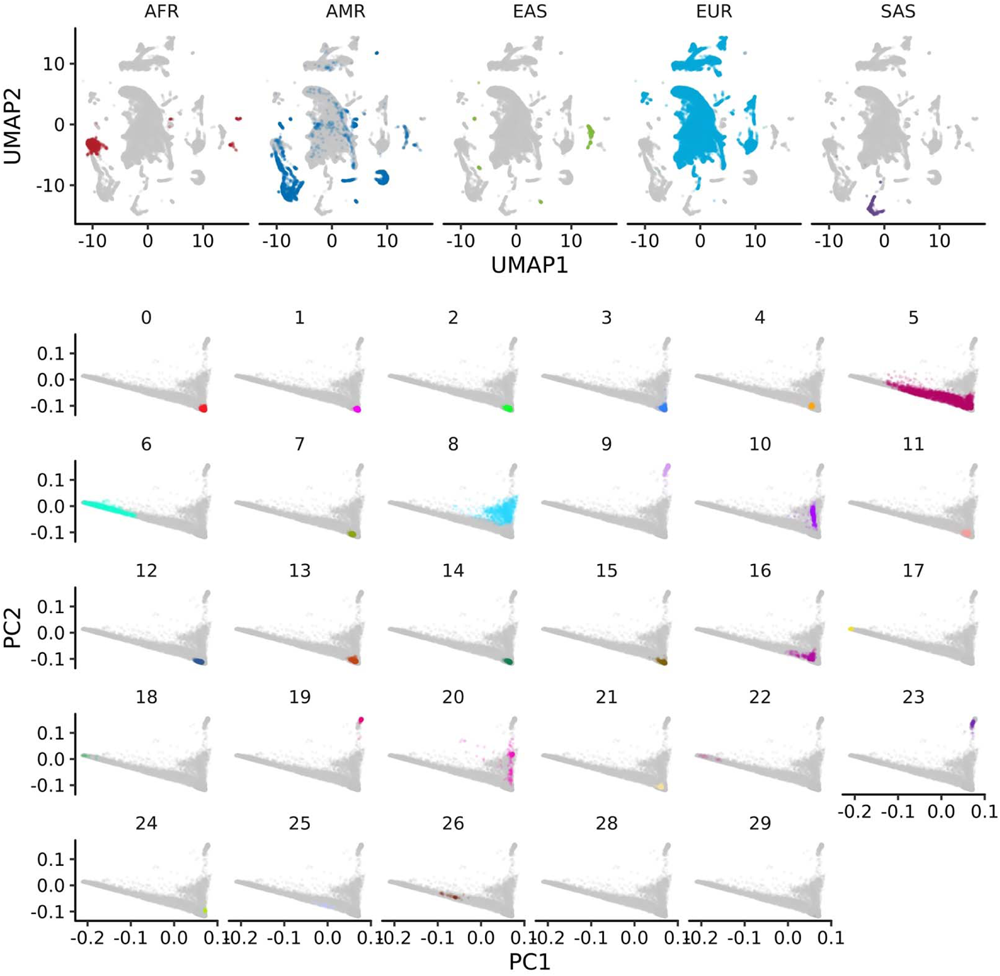
Projection of the fine genetic clusters in the continental ancestry space. **a,** The distribution of the continental ancestries in the UMAP space. Each point on the plot represents an individual participant of MGBB. To infer the continental ancestry of each participant, we utilized the K-nearest neighbor algorithm trained on the 1000 Genomes Project dataset. The colors assigned to the points represent the inferred continental ancestries. The horizontal axis corresponds to the first axis of the UMAP projection, and the vertical axis represents the second axis. This two-dimensional representation allows us to visualize the clustering and distribution patterns of different continental ancestries within the MGBB population. **b,** The distribution of the fine-scale genetic clusters in the MGBB on conventional PC space. Each point on the plot represents an individual in MGBB. The colors assigned to the points represent genetic clusters inferred by the network-based clustering method (Methods). The horizontal axis corresponds to the first genetic PC, and the vertical axis represents the second genetic PC. UMAP, Uniform Manifold Approximation and Projection; MGBB, Mass General Brigham Biobank; PC, principal component.

**Figure S2.**
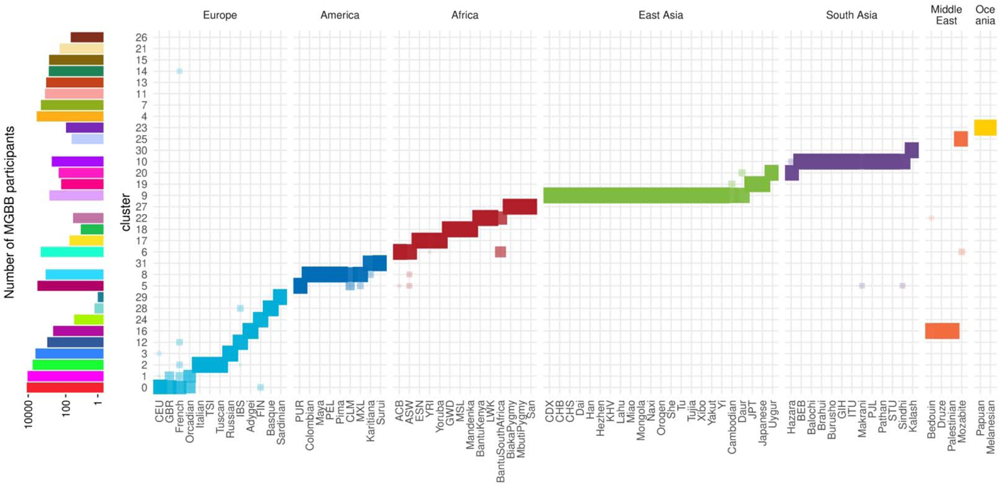
Enrichment of the reference groups in the genetic clusters inferred in MGBB. The horizontal axis shows reference populations from 1000 Genomes Project and Human Genome Diversity Project. The vertical axis shows genetic clusters inferred from MGBB. In the left panel, the heights of bars show the number of MGBB participants included in the genetic clusters. In the right panel, the size and transparency of rectangle shows intersection size between reference population and genetic clusters. The number of individuals in the clusters and the color legend are found in Supplemental Table 2. Abbreviations for reference populations will be find in Supplemental Table 7. MGBB, Mass General Brigham Biobank.

**Figure S3.**
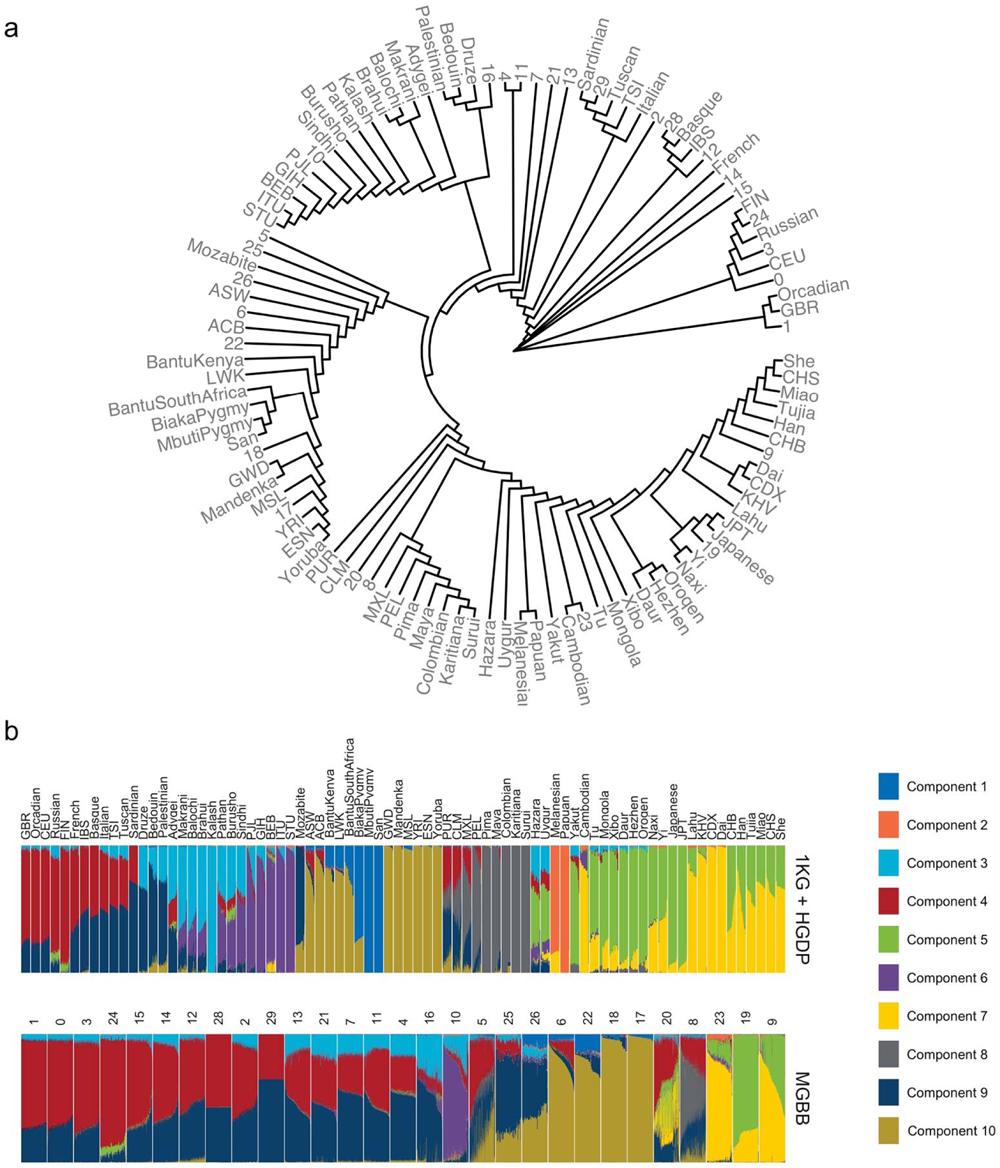
ADMIXTURE analysis for reference population (1KG + HGDP) and genetic clusters. **a,** Phylogenetic tree generated using *F*st Values. The phylogenic tree was constructed using the Neighbor-Joining method with pairwise *F*st values serving as a measure of genetic distance between populations. A higher *F*st value indicates a greater genetic differentiation between populations. The numeric numbers indicate genetic cluster in MGBB. Ancestry names indicated reference populations in 1KG or HGDP. The number indicate genetic clusters in MGBB. **b,** In each panel, a stacked column of color segments represents an individual participant. The color of each segment corresponds to one of the K = 10 ancestral components, as determined by ADMIXTURE software. The length of each colored segment within a participant’s column indicates the estimated proportion of their genome attributed to that specific ancestral population. The choice of K = 10 was informed by cross-validation results, with the aim of minimizing prediction error (Supplemental Information Figure 3). The abbreviations and detailed descriptions for the ancestral populations corresponding to each color are provided in Supplemental Table 7. 1KG, 1000 Genomes Project; HGDP, Human Genome Diversity Project; MGBB, Mass General Brigham Biobank.

**Figure S4.**
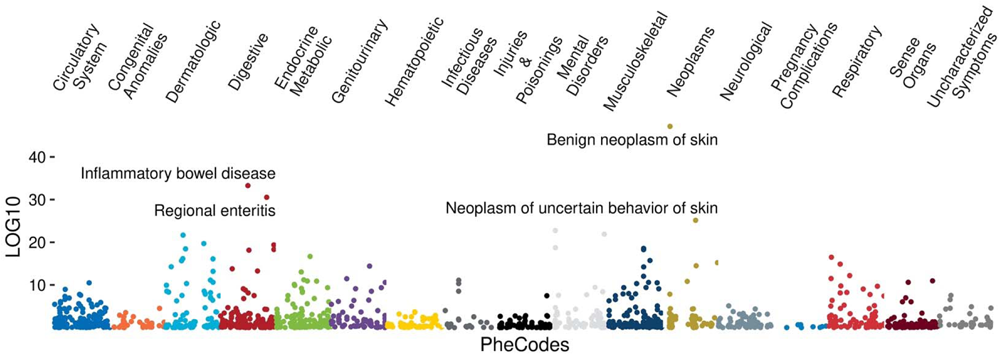
Phenome wide association analysis for cluster 4. Each dot represents the association results based on PheCodes. We examined the relationship between membership in genetic cluster 4 of the MGBB biobank and PheCodes outcomes. This association was assessed using a logistic regression model, adjusted for age and sex. MGBB, Mass General Brigham Biobank.

**Figure S5.**
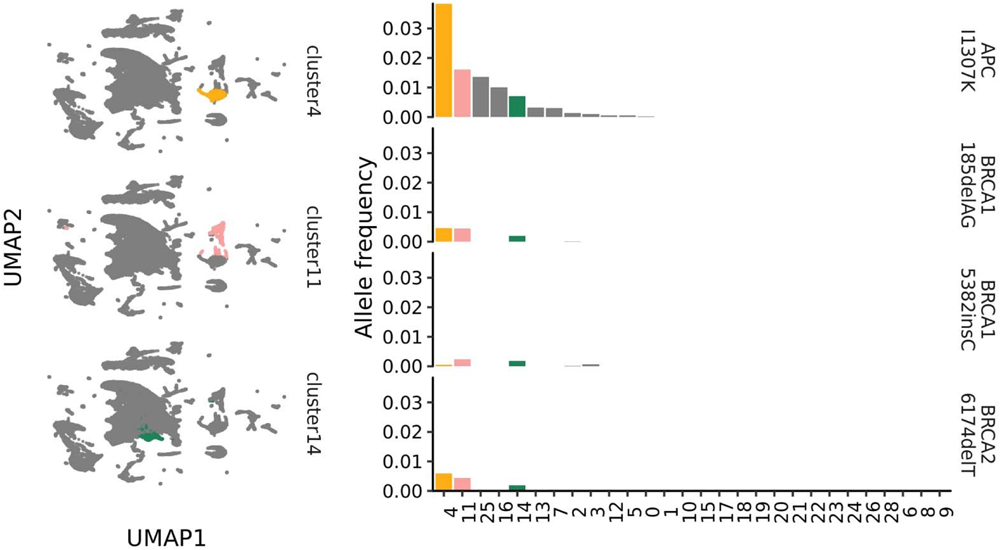
Shared founder mutations across genetic clusters 4, 11, and 14. The left panels are showing the distribution of genetic cluster 4, 11, and 14 in the UMAP space. The right panel shows the distribution of Ashkenazi founder mutations described in the previous literatures. The horizontal axes show the genetic clusters identified in MGBB, and the vertical axes show the allele frequencies determined by whole exome sequencing of MGBB by the genetic clusters. UMAP, Uniform Manifold Approximation and Projection; MGBB, Mass General Brigham Biobank.

**Figure S6.**
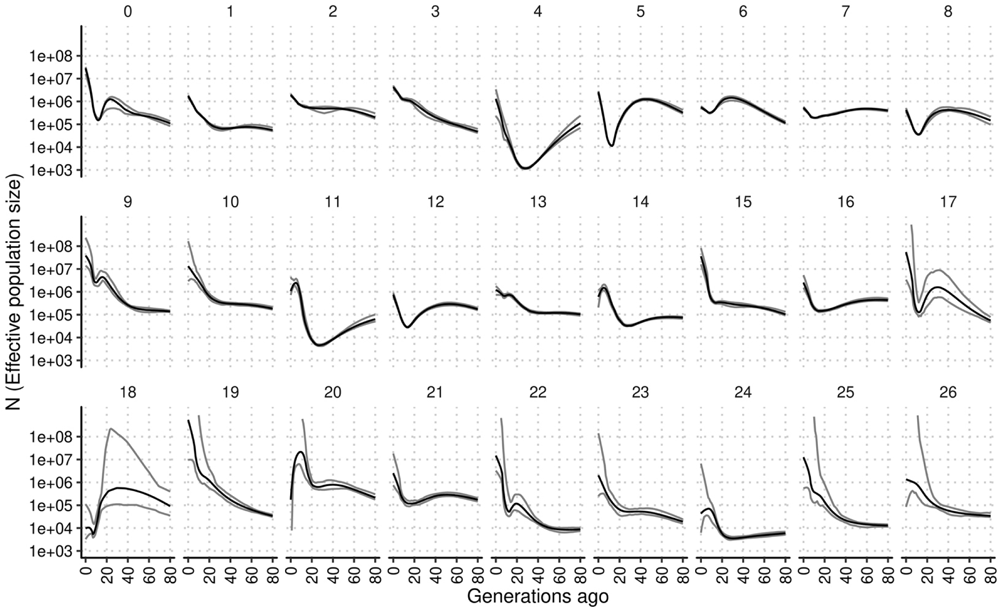
Effective population sizes in the genetic clusters in MGBB. The horizontal axes show generations ago. The vertical axes show the estimated population size. Numbers on the top of panels show the cluster identification. The black lines show the estimates and gray lines show 95% confidence interval.

**Figure S7.**
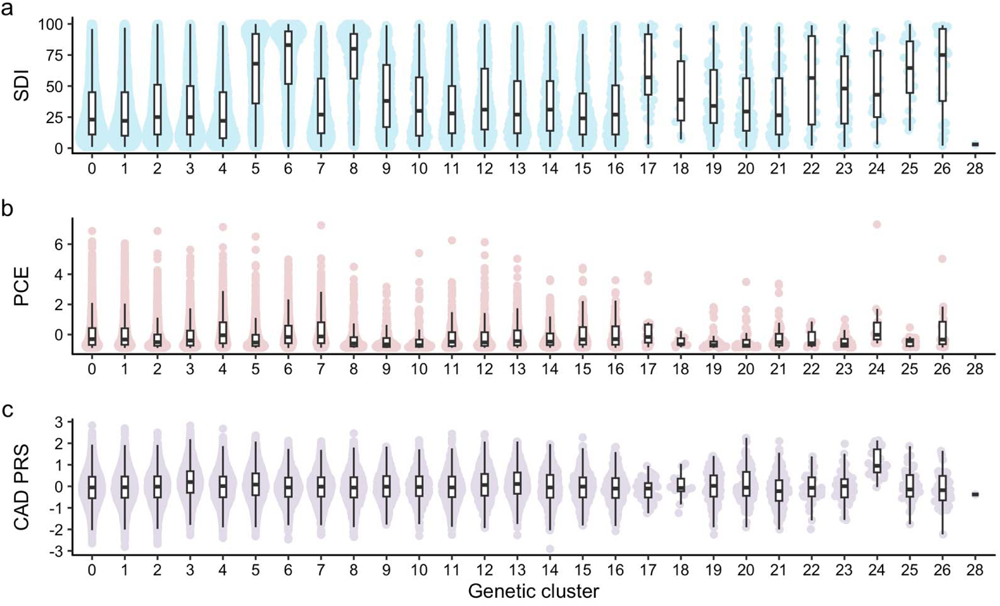
Geographical, socioeconomical, distributions of genetic clusters. Distributions of socioeconomic (**a**), clinical (**b**), and genetic risks (**c**). Each dot indicates MGBB participants. The horizontal axes show genetic cluster. SDI, social deprivation index; PCE, pooled cohort equation, CAD PRS, polygenic risk score for coronary artery disease.

**Figure S8.**
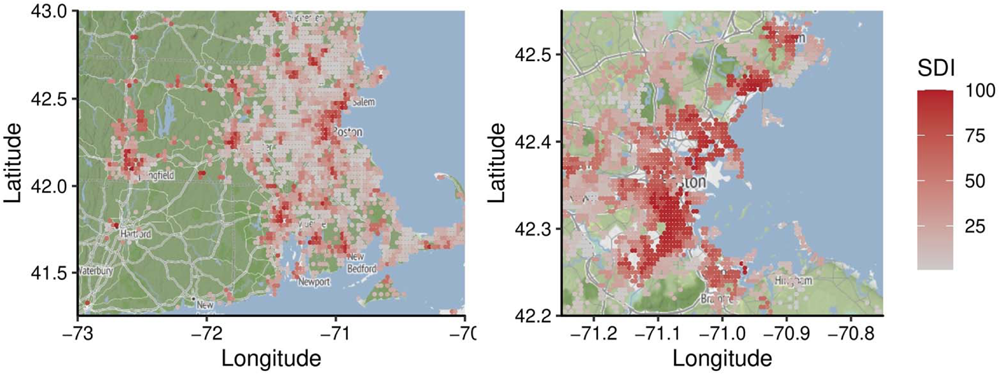
Social deprivation in Massachusetts and Greater Boston Area. The color of each grid represents the median Social Deprivation Index (SDI) of the participants from MGBB. A darker shade denotes a higher level of socioeconomic deprivation. Grids with fewer than five participants have been excluded.

**Figure S9.**
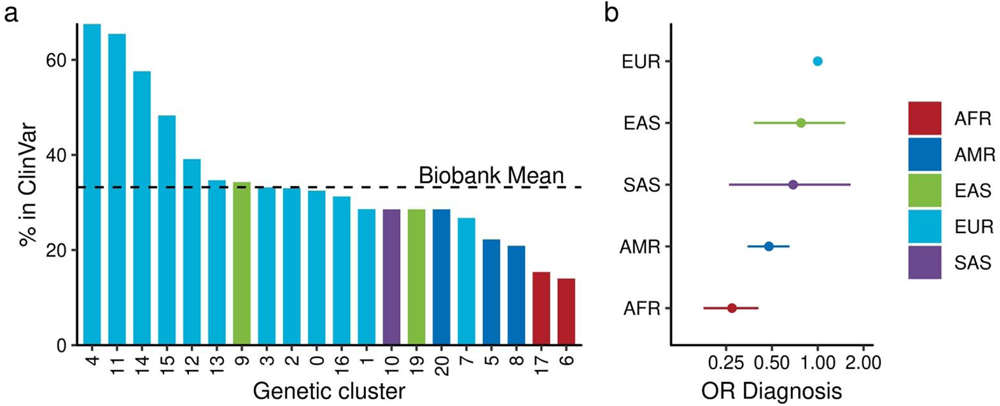
Different annotation rate for the functional variants in the pathogenic genes. **a,** The bar heights depict the proportion of pLOF variants in ACMG genes within specific genetic clusters identified in MGBB with pathogenic/likely pathogenic annotations as determined by multiple expert reviews in ClinVar. **b,** The odds ratio indicates the likelihood of pLOF variants in ACMG genes having pathogenic/likely pathogenic annotations based on multiple expert reviews. The results are presented in the continental ancestries and in reference to the European population. Error bars indicate 95% confidence intervals. OR, Odds Ratio; AFR, African; AMR, Admixed American; EAS, East Asian; EUR, European; SAS, South Asian. ACMG, American College of Medical Genetics and Genomics; MGBB, Mass General Brigham Biobank; pLOF predicted loss of function.

**Figure S10.**
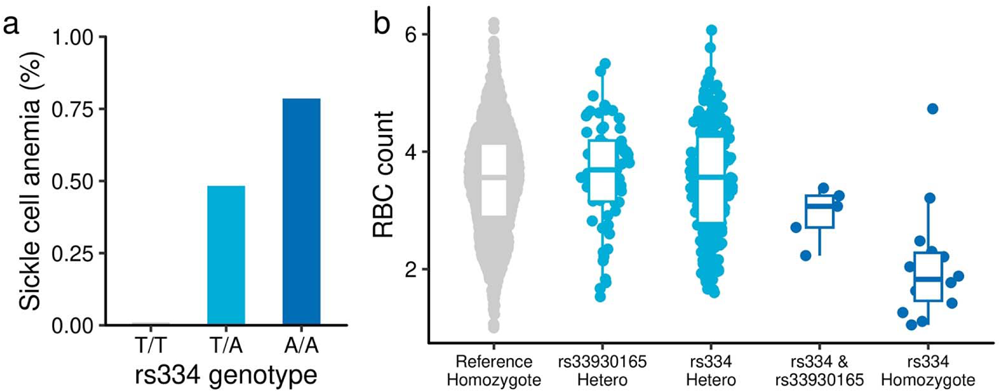
Penetrant association of rs334 for sickle cell anemia. **a,** The prevalence of Sickle cell anemia by rs334 genotypes. **b,** The RBC measurements in the MGB participants by combined genotype of rs33930165 and rs334. The box plot depicts the first and third quartiles, with the line inside the box indicating the median value. RBC, Red Blood Cell count.

