## Supplemental Information for "Decoding Genetics, Ancestry, and Geospatial Context for Precision Health"

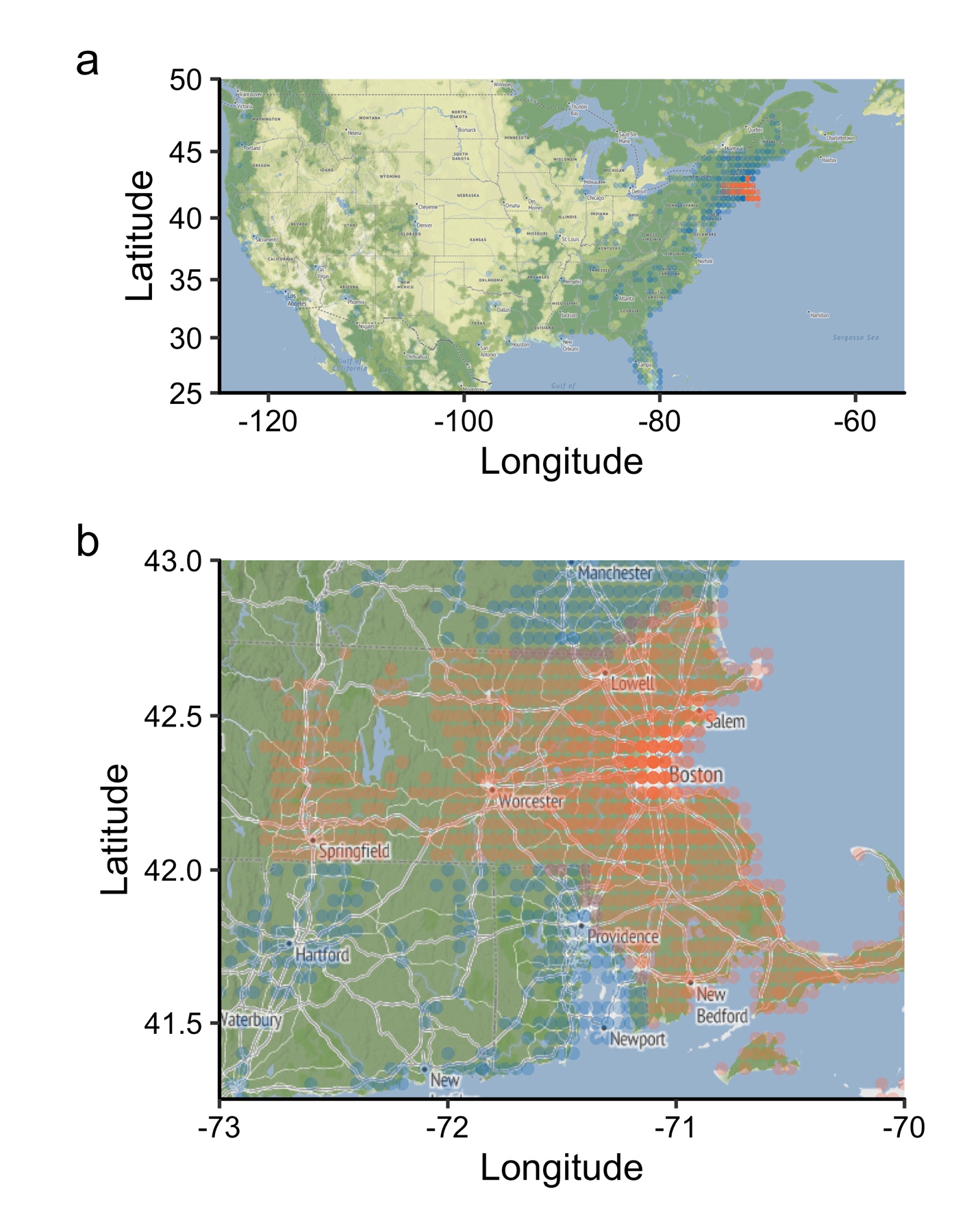

**Supplemental Information Figure 1 | Geographic distribution of MGBB participants** Each grid represents a location in the U.S. (**a**) and Massachusetts (**b**) with a minimum of 5 participants. Grids in Massachusetts are colored orange, while grids outside of Massachusetts are blue. The transparency of each grid indicates its population density, with denser areas being less transparent. MA, Massachusetts.

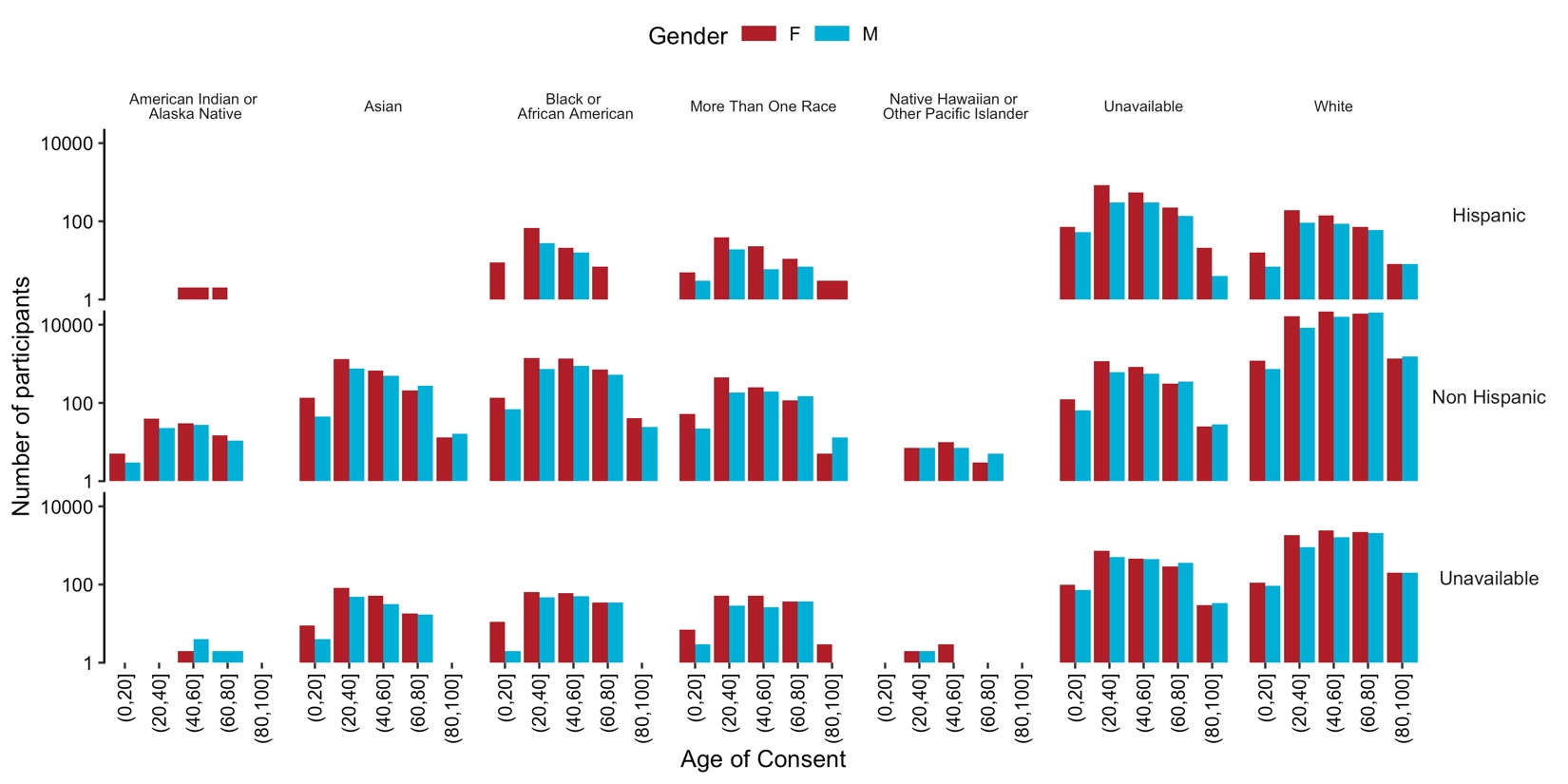

**Supplemental Information Figure 2 | Self-reported Race and Ancestry in MGBB** The height of each bar represents the number of participants in that category. Bar colors denote self-reported gender. Vertical categories correspond to self-reported ethnicity, while horizontal categories refer to self-reported race. MGBB, Mass General Brigham Biobank.

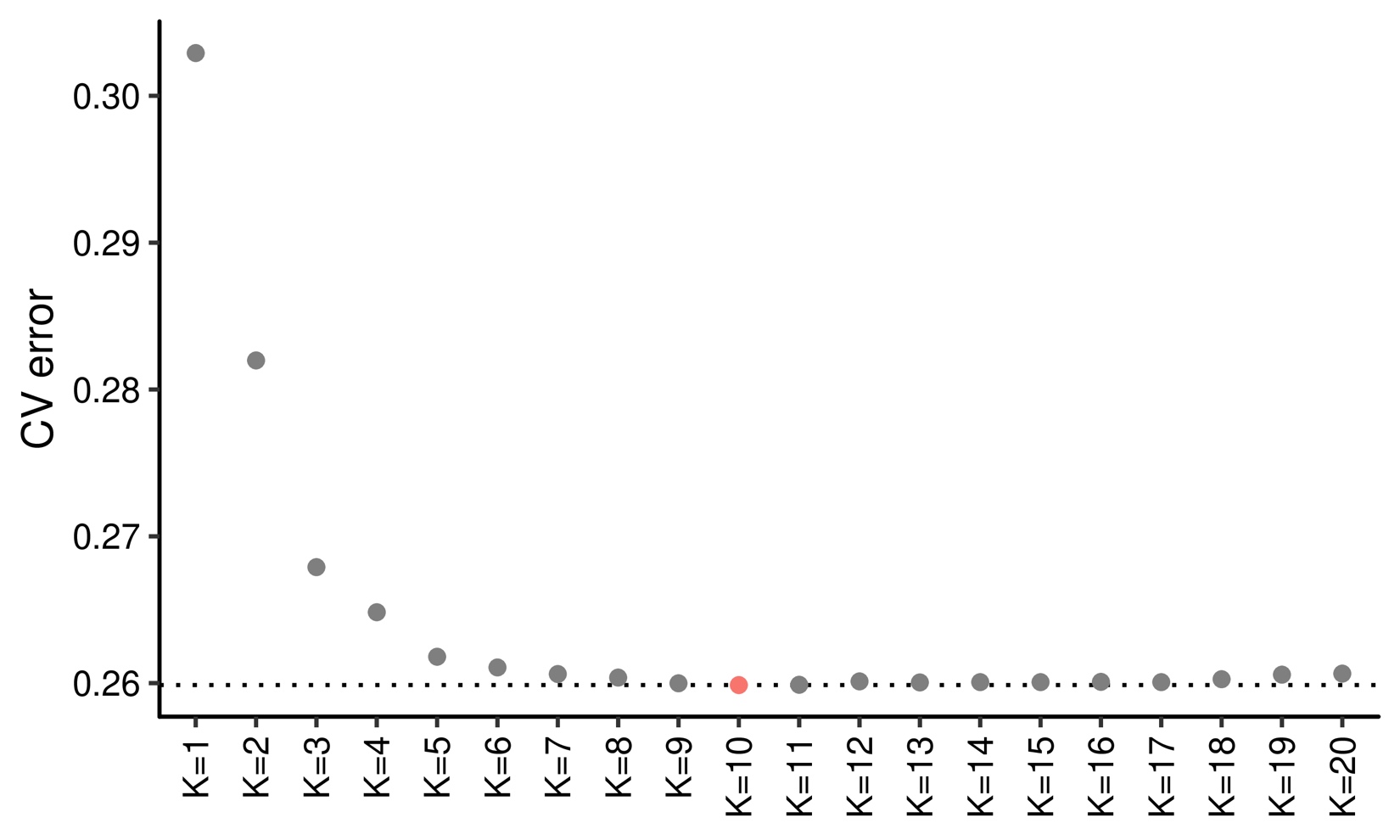

**Supplemental Information Figure 3 | Cross validation in ADMIXTURE analysis** The horizontal axis shows parameter K used in the cross-validation experiments. The vertical axis shows error in cross-validation (CV). K = 10 showed the least CV error.

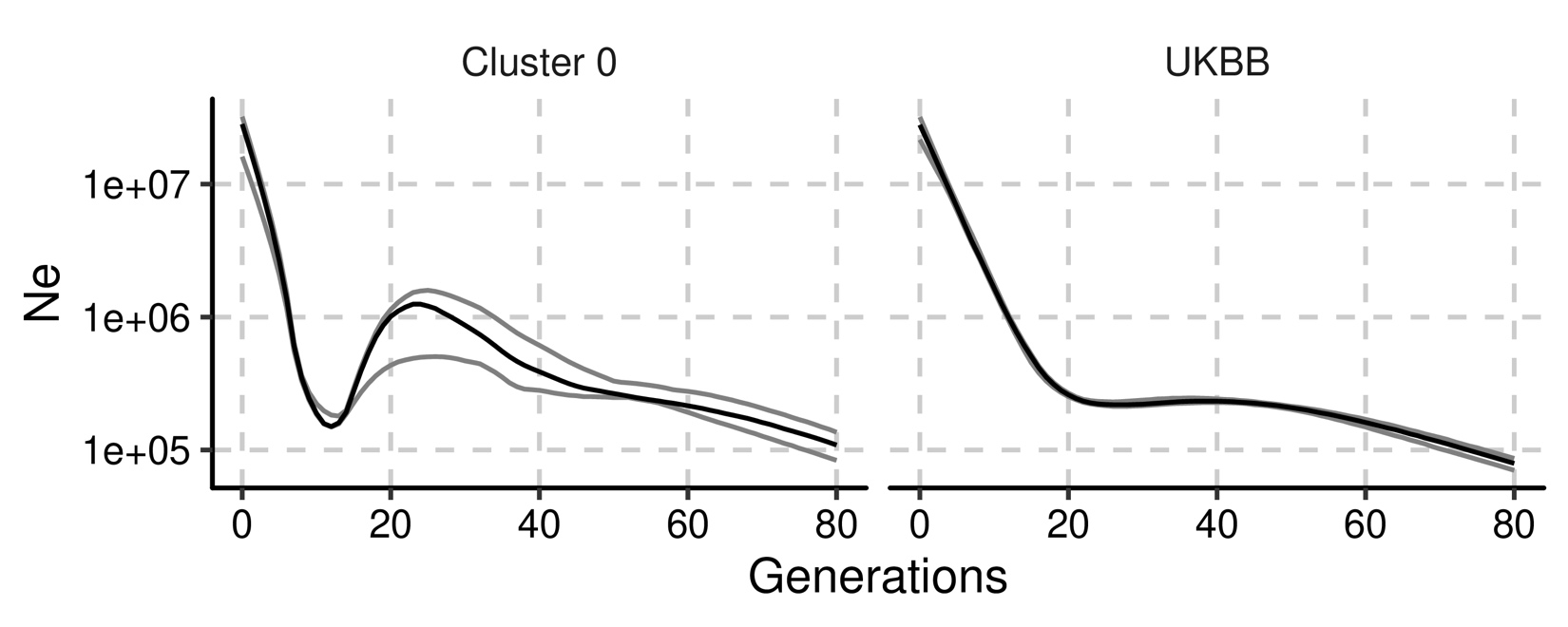

**Supplemental Information Figure 4 | Number of effective population size in British cluster in Massachusetts and United Kingdom** The horizontal axes shows the generations ago from current generation. Cluster 0 indicates central European like cluster in MGBB and UKBB indicates white population in the UKBB. Ne was estimated from the phased genotypes in the same sample sizes. The black lines show the estimated Ne and gray lines show its 95% confidence interval. Ne, Number of effective population size. UKBB, UK biobank; MGBB, Mass General Brigham Biobank.

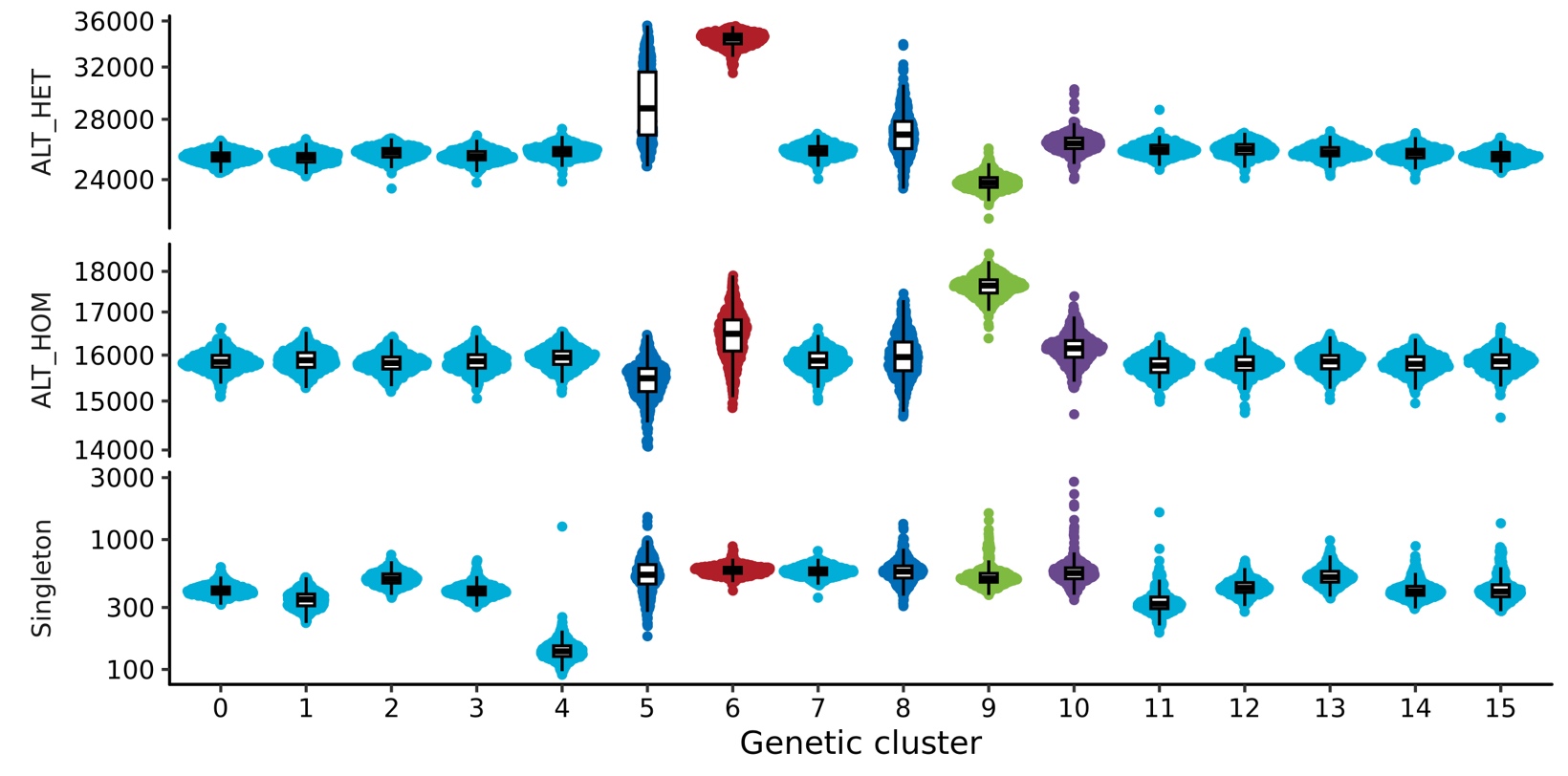

**Supplemental Information Figure 5 | The genetics of ancestral clusters** Each dot indicates exome sequenced individuals grouped by genetic ancestries inferred in MGBB. The distribution was re-estimated by random sampling 500 individuals in each cluster. The colors indicate continental ancestries dominantly observed in each genetic cluster. ALT_HET, heterozygote alternate allele counts; ALT_HOM, homozygote alternate allele counts.

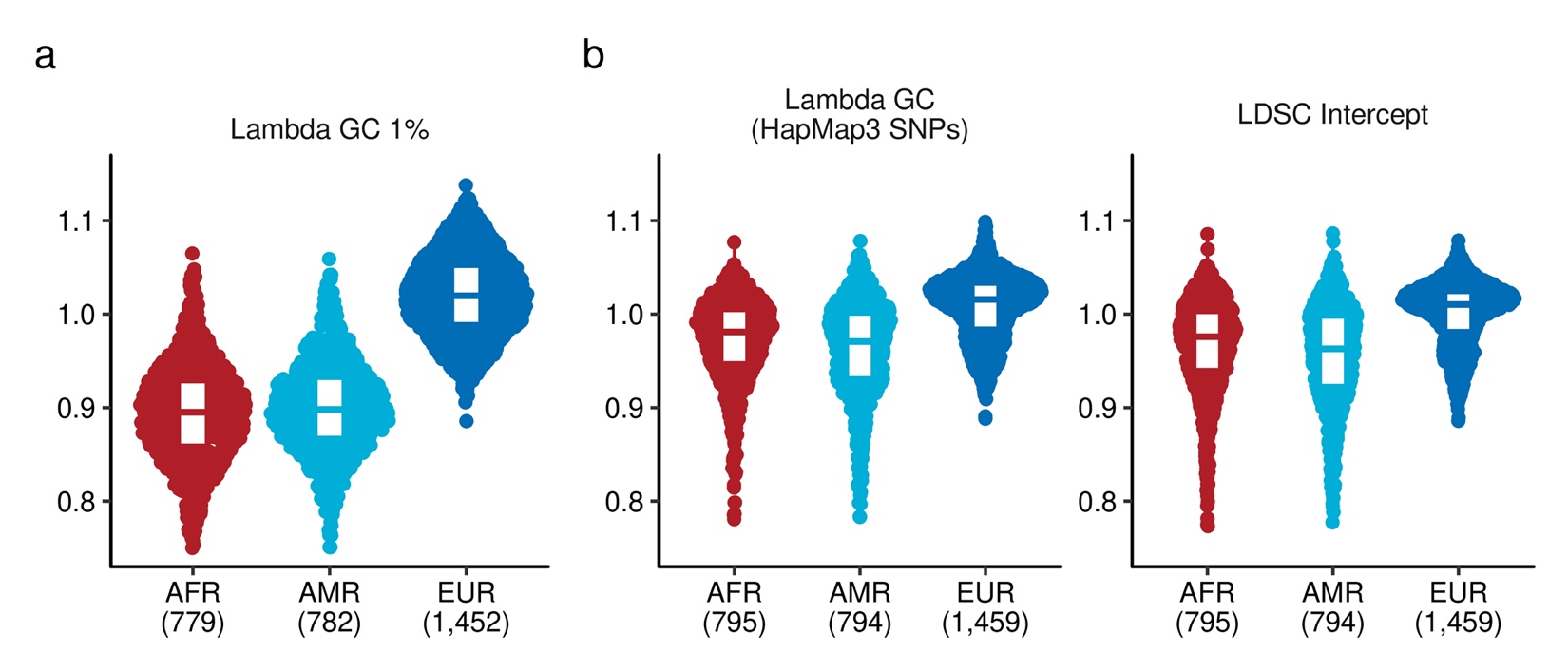

**Supplemental Information Figure 6 | Inflation statistics in Rare and Common Variant PheWAS**

**a,** Lambda GC at top 1 percentile of the test statistics in rare variant burden association testing. **b,** Lambda GC and LDSC Intercept in the common variant association testing. Lambda GC was computed using Hapmap3 SNPs. The numbers in the parathesis indicates the number of tested PheCodes in the designated ancestry group. LDSC, Linkage Disequilibrium Score regression.

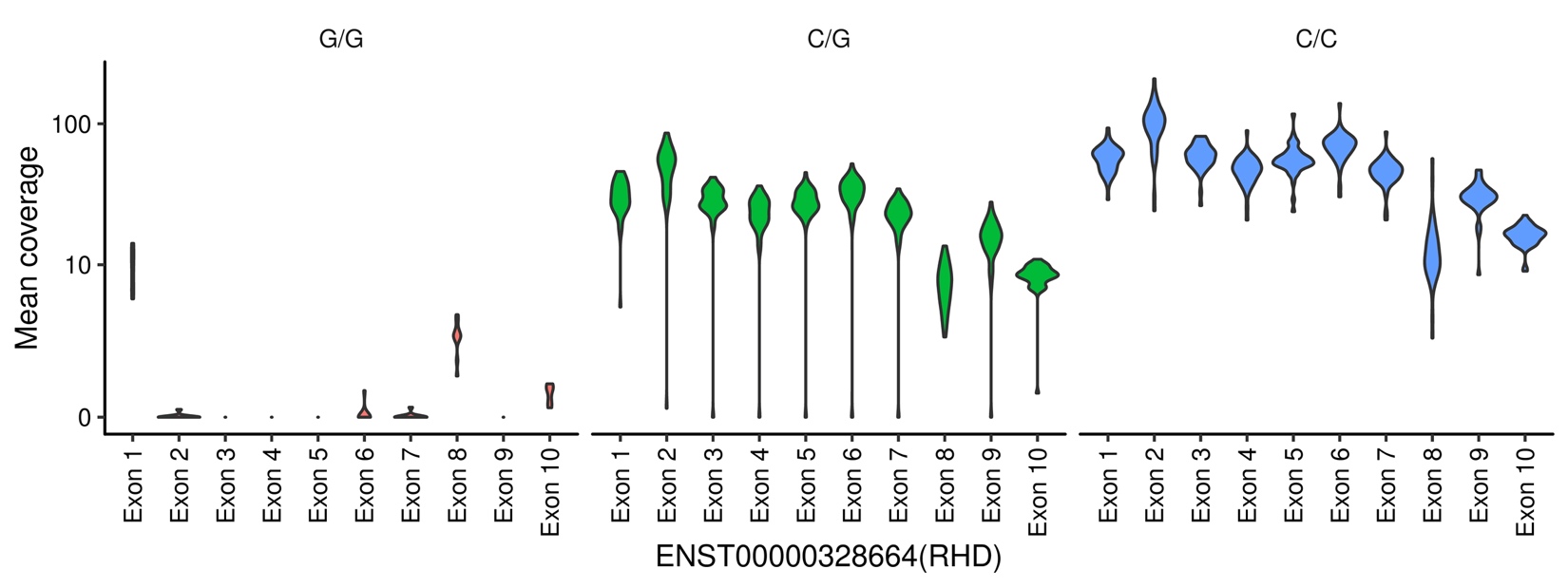

**Supplemental Information Figure 7 | RHD deletion tagged by rs72660908** We computed the mean coverage of exons of RHD gene for randomly sampled 100 participants of MGBB sequenced by whole exome sequence. The vertical axis shows the distribution of mean coverages of each exon. The data was presented by hard-called imputed genotypes of rs72660908.

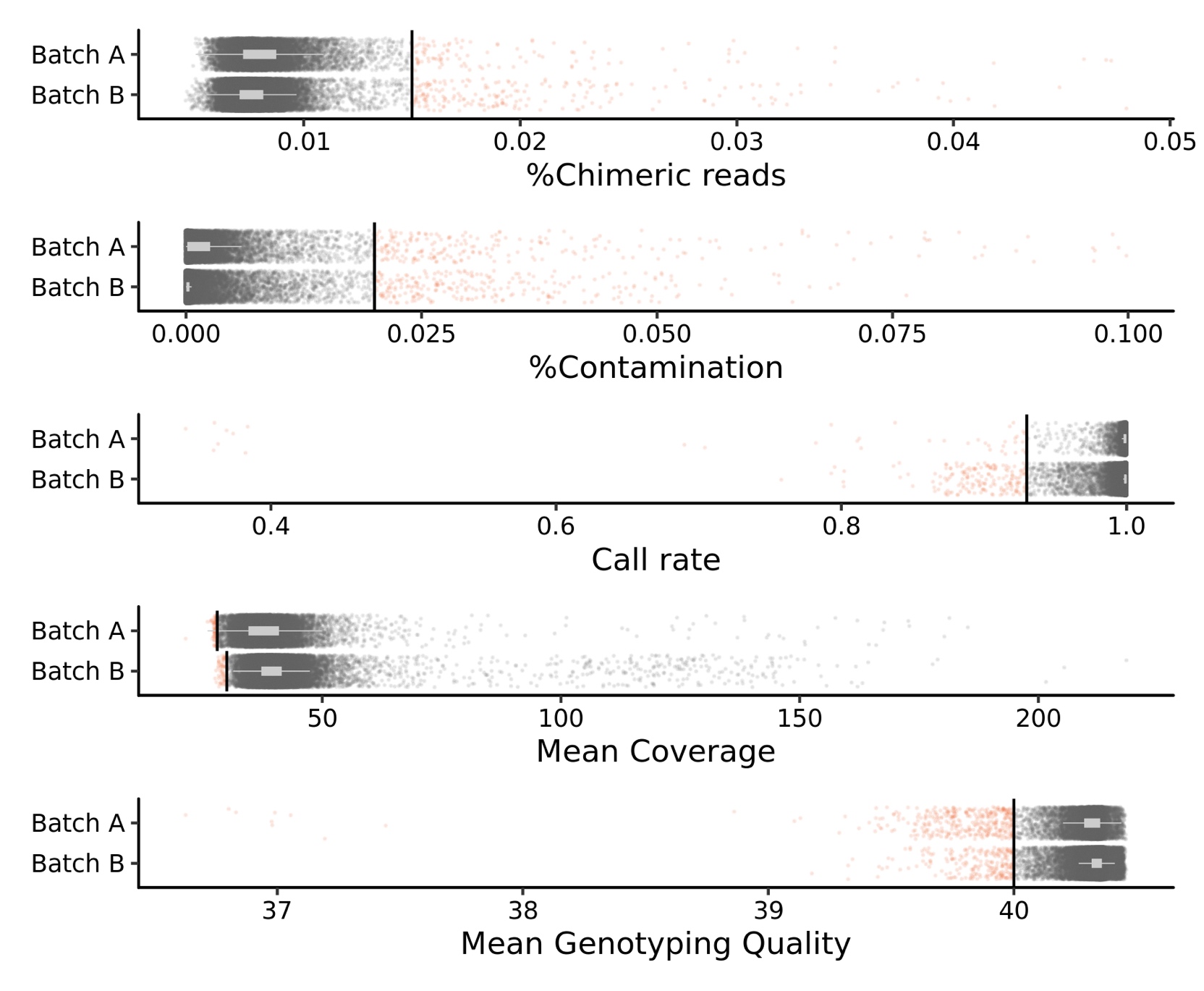

**Supplemental Information Figure 8 | Hard filtering for sample QC in MGBB exome data**

Each dot represents an individual. Gray dots indicate samples that have passed quality control, while red dots indicate samples that have not. Batch A comprises 13,331 samples, while Batch B contains 40,089 samples.

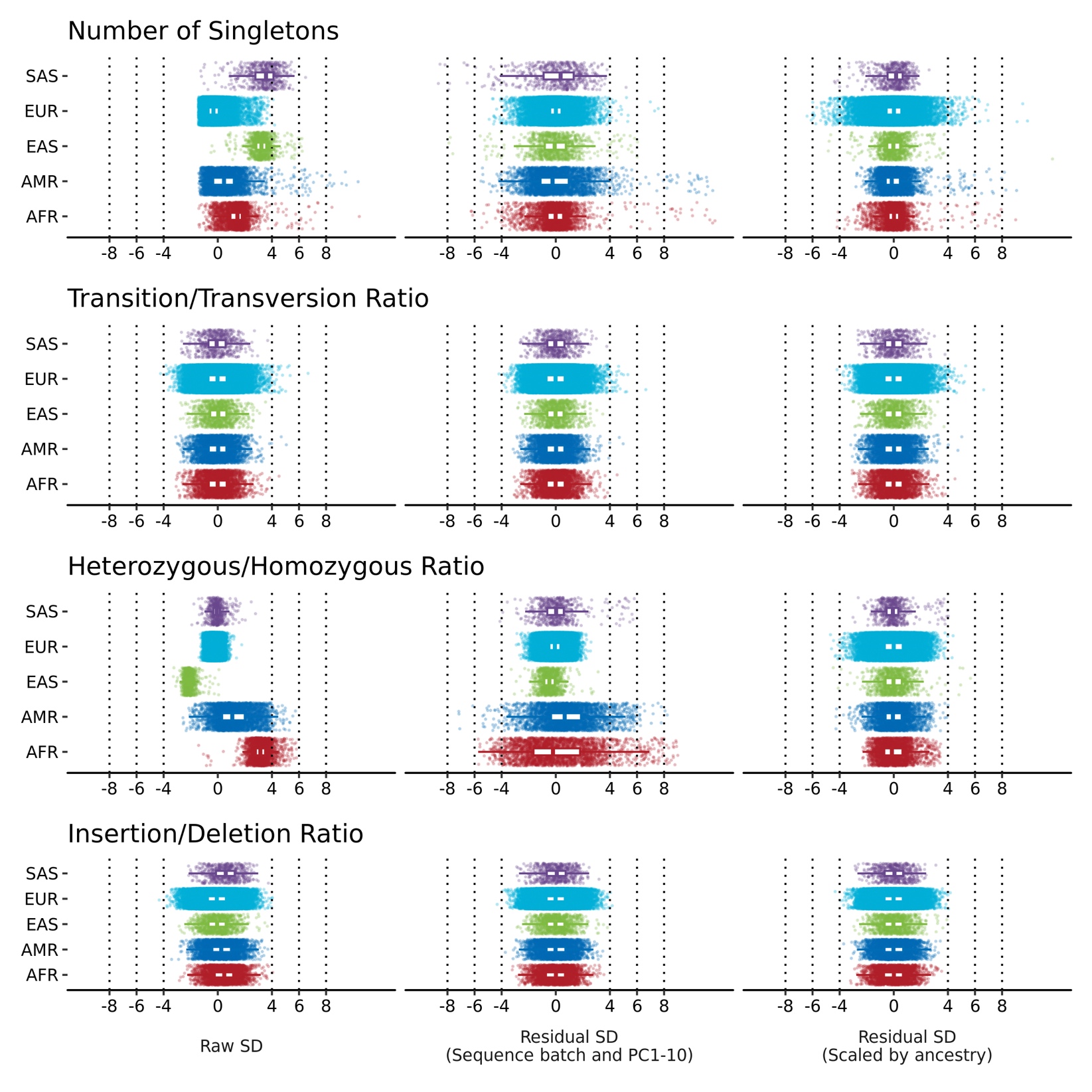

**Supplemental Information Figure 9 | Soft filtering for sample QC in MGBB exome data**

Each point represents an individual, with the color denoting their continental ancestry. The QC metrics are linearly scaled (on the left), residualized using the first ten genetic principal components (in the middle) and scaled by ancestry (on the right).
